# Spatiotemporal evolution of the ccRCC microenvironment links intra-tumoral heterogeneity to immune escape

**DOI:** 10.1101/2022.07.11.22277322

**Authors:** Mahdi Golkaram, Fengshen Kuo, Sounak Gupta, Maria I. Carlo, Michael L. Salmans, Raakhee Vijayaraghavan, Cerise Tang, Vlad Makarov, Phillip Rappold, Kyle A. Blum, Chen Zhao, Rami Mehio, Shile Zhang, Jim Godsey, Traci Pawlowski, Renzo G. DiNatale, Luc GT Morris, Jeremy Durack, Paul Russo, Ritesh R. Kotecha, Jonathan Coleman, Ying-Bei Chen, Victor E Reuter, Robert J Motzer, Martin H. Voss, Li Liu, Ed Reznik, Timothy A. Chan, A. Ari Hakimi

## Abstract

**Background:** Intratumoral heterogeneity (ITH) is a hallmark of clear cell renal cell carcinoma (ccRCC) that reflects the trajectory of evolution and influences clinical prognosis. Here we seek to elucidate how ITH and tumor evolution during immune checkpoint inhibitor (ICI) treatment can lead to therapy resistance.

**Methods:** Here, we spatiotemporally profiled the genomic and immunophenotypic characteristics of 29 ccRCC patients, including pre- and post-therapy samples from 17 ICI treated patients. Deep multi-regional whole exome and transcriptome sequencing were performed on 29 patients at different time points before and after ICI therapy. T cell repertoire was also monitored from tissue and peripheral blood collected from a subset of patients to study T cell clonal expansion during ICI therapy.

**Results:** Angiogenesis, lymphocytic infiltration, and myeloid infiltration varied significantly across regions of the same patient, potentially confounding their utility as biomarkers of ICI response. Elevated ITH associated with a constellation of both genomic features (HLA LOH, CDKN2A/B loss) and microenvironmental features, including elevated myeloid expression, reduced peripheral T cell receptor (TCR) diversity, and putative neoantigen depletion. Hypothesizing that ITH may itself play a role in shaping ICI response, we derived a transcriptomic signature associated with neoantigen depletion that strongly associated with response to ICI and targeted therapy treatment in several independent clinical trial cohorts.

**Conclusions:** These results argue that genetic and immune heterogeneity jointly co-evolve and influence response to ICI in ccRCC.

**Trial registration:** We completed a single-arm pilot study at Memorial Sloan Kettering Cancer Center (MSKCC; ClinicalTrials.gov identifier NCT02595918) to examine the safety and feasibility of neoadjuvant nivolumab in patients with localized RCC.

## Background

Clear cell renal cell carcinoma (ccRCC) is the most common histological subtype of kidney cancer and demonstrates a high response rate to immune checkpoint inhibitors such as nivolumab, pembrolizumab and ipilimumab [1–3]. However, only a subset of ccRCC patients respond to ICI, and biomarkers for ICI response in other disease settings such as tumor mutation burden, neoantigen load and mismatch repair deficiency do not associate with ICI response in ccRCC [4–7]. Recently, several studies have identified transcriptomic microenvironmental features including angiogenic gene expression, T-cell infiltration, and myeloid activation that correlate with response or resistance to ICI and combination therapies in ccRCC [7–13]. This suggests that the ccRCC microenvironment, in addition to genomic factors, influences ICI response.

In parallel, recent work has demonstrated the prevalence of ITH in untreated ccRCC [14]. This study has largely focused on heterogeneity in the presence of key driver mutations and copy number alterations and have demonstrated that ccRCC tumors follow one of a small number of evolutionary trajectories, each of which are associated with distinct patterns of genomic ITH and clinical prognosis. However, the potential for non-genomic heterogeneity in the tumor microenvironment, including but not limited to variability in the amount and identity of immune cells in spatially distinct regions of the same tumor is overlooked. Recently, we and others described substantial heterogeneity in the tumor-microenvironment (TME) in several small cohorts of ccRCC tumors both in the treatment-naïve and treatment-exposed settings, raising the possibility that heterogeneity in the TME may itself shape the evolution of the tumor and its likelihood to respond to therapy [15, 16].

In this study, we hypothesized ccRCC tumors with elevated ITH constitute a genomically and immunologically distinct class of tumors, with distinguishing clonal/subclonal genomic alterations, immunologic profiles, and therapeutic response trajectories. To test this hypothesis, we utilize whole exome sequencing (WES), whole transcriptome sequencing (WTS), TCRseq, and histopathologic multi-regional data across a cohort of untreated and ICI exposed patients from a phase 2 clinical trial to reveal the molecular determinants of therapy response in ccRCC (Fig. 1 and Table S1). Our integrated analysis demonstrated that ITH is highly correlated among genomic, transcriptomic, and TME characteristics. ITH-high tumors are enriched for features including SETD2 and PBRM1 mutations, HLA loss of heterozygosity (HLA LOH), and CDKN2A/B loss. Immunologically, ITH-high tumors display a depletion of putative neoantigens, elevated myeloid activation, and reduced T cell diversity, that are in aggregate associated with escape from the anti-tumor immune response. Premised on these observations, we developed a transcriptional signature for immune escape which correlates with distinct histopathologic patterns and is associated with ICI resistance across several diverse clinical trial cohorts.

**Figure 1.**
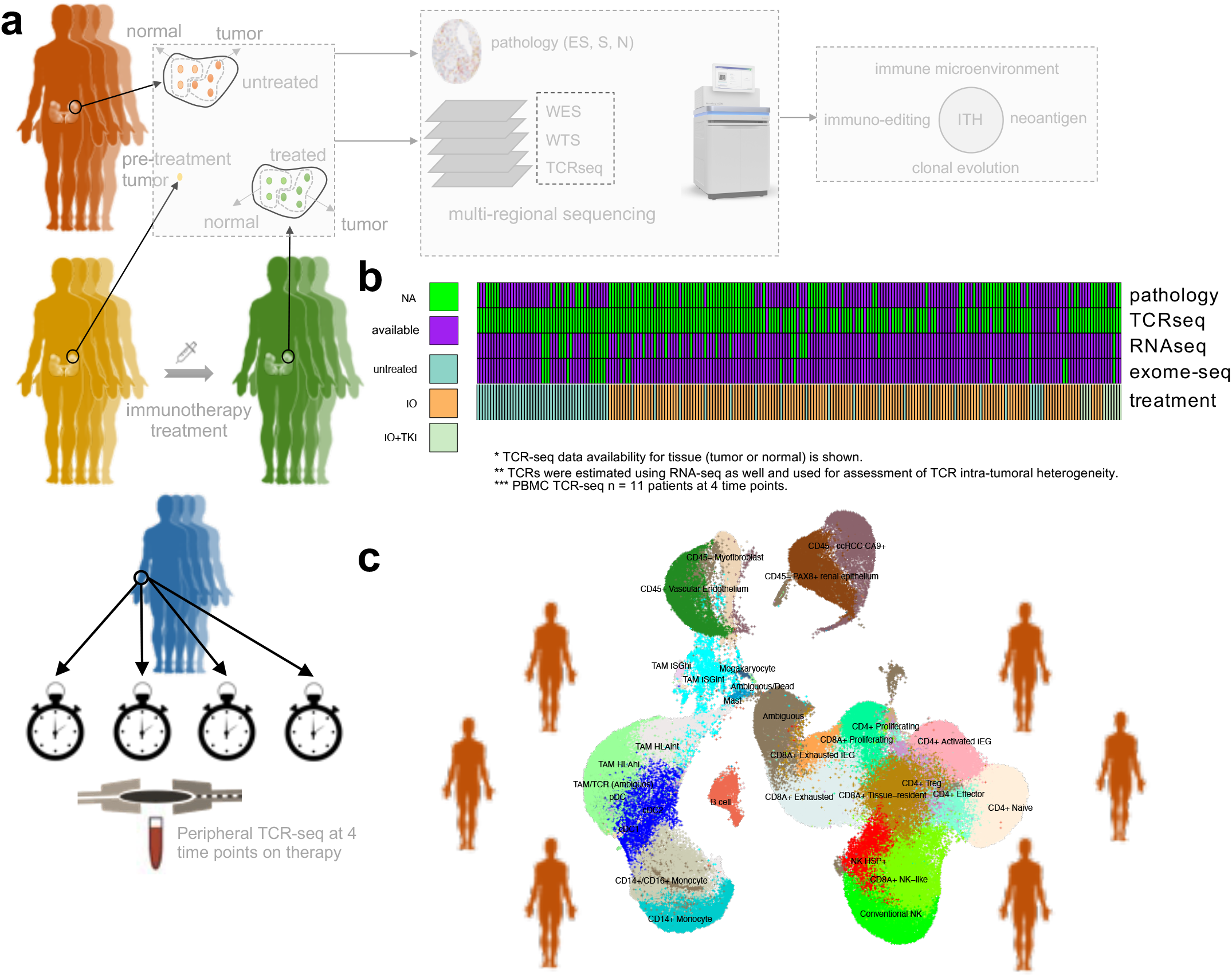
Patient characteristics and study design. **a)** Multi-regional multi-omics was performed on 29 patients. **b)** In total, 6 out of 29 patients were untreated and the rest were treated with ICI or in combination with TKI. TCRseq of PBMC was performed at 4 time points on therapy for a subset of patients. In addition, pathological review was performed to assign N-TIL (tumors sparsely infiltrated by TILs), S-TIL (tumors dominated by stromal TILs), and ES-TIL (tumors with substantial levels of both epithelial and stromal TILs) classes to a subset of patients. **c)** Additionally, scRNAseq data for 6 patients were available from [15].

## Materials and Methods

### Sample acquisition

After acquiring informed consent and institutional review board approval from Memorial Sloan Kettering Cancer Center (MSK), partial or radical nephrectomies were performed at MSK (New York) and stored at the MSK Translational Kidney Research Program (TKRCP). Samples were flash frozen and stored at −80 degrees Celsius prior to molecular characterization. Clinical metadata was recorded for all tumor samples. All patients represent clear cell histology and were treated via ICI alone or in combination with tyrosine kinase inhibitor (TKI). All treatments were administered prior to surgery in a neo-adjuvant setting and biopsies were collected. Detailed clinical data and treatment regimen for each patient is included in Table S2.

### Untreated cohort

Using and institutional database we identified six patients with advanced or metastatic ccRCC that underwent nephrectomy with multiregional data available MR01,02,03,05,06, SC03. Clinical and pathologic data is available in Table S2.

### Neoadjuvant multiregional cohort

This open-label, single-arm, pilot study was done at Memorial Sloan Kettering Cancer Center and funded through the National Institute of Health’s Cancer Therapy Evaluation Program (CTEP). Patients received nivolumab (dose initially 3 mg/kg, then protocol amended to 240 mg flat dose) every 2 weeks for 4 treatments. Surgery was planned 7-14 days after the last dose. Prior to starting therapy, all patients had a kidney biopsy to confirm ccRCC, and tumor staging with renal protocol MRI and CT of the chest. After 4 doses and prior to surgery, patients also had a renal protocol MRI. Changes in primary tumor size were assessed according to Response Evaluation Criteria in Solid Tumors (RECIST) version 1.1. Resection of the primary tumor and lymph nodes was done according to standard institutional procedures. From May 27, 2016, to September 9, 2019, 21 patients were screened and 18 were enrolled into the study of which 17 had available genomic data. Baseline patient characteristics are in Supplementary Table S2. All patients had localized disease at time of enrollment and biopsy-proven clear cell RCC. Perioperative and pathological details are included in Supplementary Table S2. Median time to nephrectomy after the last dose of nivolumab was 10.5 days (range, 9-13 days).

### Metastatic multiregional Cohort

Using an institutional database, we identified 6 additional patients who had received ICI prior to nephrectomy (Supplementary Table S2). All patients had metastatic disease at time of ICI; two received anti-VEGF therapies before ICI.

### Multi-regional sampling

For the prospective neoadjuvant trial and the “MR” samples single region biopsies were obtained preoperatively. Following nephrectomy, tumor were bivalved and 5 regions were chosen: One region from the tumor center and 4 from each quadrant (upper medial, upper later, lower medial, lower lateral). Grossly necrotic or hemorrhagic regions were avoided. For the remaining samples (those treated with definitive immunotherapy “SC”) regions were taken from distinct regions of tumors separated by 1-2 cm avoiding grossly necrotic or hemorrhagic regions).

### Whole exome sequencing

Libraries for whole exome sequencing were generated with TruSight Oncology DNA Library Prep Kit with 40ng input DNA per sample. TruSight Oncology index PCR products were directly used for enrichment and target exome enrichment was performed using the IDT xGen Universal Blockers and IDT xGen Exome Research panel. A single-plex hybridization was done overnight at 65°C. Accuclear dsDNA Ultra High Sensitivity assay (Biotium) was used for library quantification of the post-enriched libraries. Post enrichment libraries were normalized using bead-based normalization and pooled. Samples were sequenced with 101 bp paired-end reads on Illumina NovaSeq™ 6000 S4 flow cell using the XP workflow for individual lane loading (12-plex per lane). On average, each sample yielded 500 million reads and MEDIAN_TARGET_COVERAGE depth of 360X.

### Whole transcriptome sequencing

Libraries for whole transcriptome RNA-seq were generated with Illumina TruSeq Stranded Total RNA. 100 ng RNA was used as input for Ribo-Zero rRNA Removal Kit, with Illumina TruSeq RNA UD Indexes (96 indexes) for sample indexing. Qubit dsDNA High Sensitivity assay (Thermo Fisher Scientific) was used for library quantification. Sequencing was done on Illumina NovaSeq™ 6000 S2 (36-plex) or S4 (72-plex) flow cell with 76 bp paired-end sequencing to produce ∼200 million paired reads per library.

### T-cell repertoire sequencing

Libraries for T-cell repertoire sequencing were generated with AmpliSeq for Illumina Library PLUS paired with AmpliSeq cDNA Synthesis for Illumina with 100 ng RNA input per cDNA synthesis reaction. The TCR beta-SR Panel was used for generating amplicons, and AmpliSeq CD Indexes Set A for Illumina were used for sample barcodes. Qubit dsDNA High Sensitivity assay (Thermo Fisher Scientific) was used for library quantification. Sequencing was done on the NextSeq 550 (41-plex) with 151 bp paired-end sequencing to produce ∼5 million paired reads per library.

### WTS pipeline

WTS raw read sequences were aligned against human genome assembly hg19 by STAR 2-pass alignment [17]. QC metrics, for example general sequencing statistics, gene feature and body coverage, were then calculated based on the alignment result through RSeQC. WTS gene level count values were computed by using the R package GenomicAlignments [18] over aligned reads with UCSC KnownGene [19] in hg19 as the base gene model. The union counting mode was used and only mapped paired reads after alignment quality filtering were considered. Finally, gene level FPKM (Fragments Per Kilobase Million) and raw read count values were computed by the R package DESeq2 [20].

### ESTIMATE

The ESTIMATEScore, which is the estimate of the presence of stromal and immune cells in tumor tissue, is calculated through the ESTIMATE R package [21] based on a given gene expression profile in FPKM.

### Immune deconvolution analysis

Two distinct popular computational methods, ssGSEA [22] and CIBERSORT [23], were chosen for immune deconvolution analysis. Signature gene lists of immune cell types for ssGSEA were obtained from Bindea et al. [24] and Senbabaoglu et al. [3]. ssGSEA takes the sample FPKM WTS expression values as the input and computes an enrichment score for the given gene list of immune cell type relative to all other genes in the transcriptome. On the other hand, CIBERSORT also takes FPKM WTS expression values as the input but uses a signature gene expression matrix of interest immune cell types instead to compute the infiltration level of each immune cell type. The LM22 immune cell signature which was validated and published along with CIBERSORT is used. We also used FRICTION [25] to deconvolute WTS into absolute CD8 and CD4 T cells as well as CD19 B cells.

### HERV quantification

We used WTS to quantify HERVs as described before [25]. Briefly, all WTS reads were aligned (using STAR aligner with optimized multi mapping options) to a custom genome built were human reference (hg19) and HERV specific reference are combined. Then reads aligned to non-HERV genes are removed and the rest are annotated. 3 samples contained super high median HERVs (Grubbs test P<0.05) and removed for better visualization.

### WES analysis pipeline

Raw sequencing data were aligned to the hg19 genome build using the Burrows-Wheeler Aligner (BWA) version 0.7.17 [26]. Further indel realignment, base-quality score recalibration and duplicate-read removal were performed using the Genome Analysis Toolkit (GATK) version 3.8 [27] following raw reads alignments guidelines [28]. VarScan 2 [29], Strelka v2.9.10 [30], Platypus 0.8.1 [31], Mutect2 – part of GATK 4.1.4.1 [28], Somatic Sniper version 1.0.5.0 (SNVs only), and [32] were used for small variant calling and combination of 2 out 5 callers are reported as per Cancer Genome Atlas Research Network recommendations [33]. Variants were filtered using the following criteria:

1. Tcov > 10 & Taf >=0.04 & Ncov > 7 & Naf <= 0.01 & Tac > 4 are set to Pass
2. Common SNPs are eliminated by comparison to snp142.vcf
3. Rare variants found in dbSNP are kept if Naf = 0
4. Variants with Tcov < 20 or Tac < 4 are marked as low_confidence
5. Only variants called by more than 1 caller are reported.
6. Common variables gnomAD v 2.1.1 are excluded.

Variants were annotated using Ensembl Variant Effect Predictor (VEP) [34]. Additional optimization and filtering are applied for INDELS. INDELS in blacklisted regions (https://www.encodeproject.org/annotations/ENCSR636HFF/) and low mappability regions (such as repeat maskers) are excluded as per [35]. Combination of filtered SNV and INDELS are used by maftools R package is used to generate oncoplots and summary plots, as per author’s recommendations https://www.bioconductor.org/packages/release/bioc/vignettes/maftools/inst/doc/maftools.html

All nonsynonymous point mutations identified as above were translated into strings of 17 amino acids with the mutant amino acid situated centrally using a bioinformatics tool called NAseek. A sliding window method is used to identify the 8-11 amino acid substrings within the mutant 17-mer that had a predicted MHC Class I binding affinity of ≤ 2 %Rank to one (or more) of the patient-specific HLA alleles. Binding affinity for the mutant and corresponding wild type nonamer is analyzed using NetMHCpan4.0 software. Only neoantigens with a TPM>1 are considered to be expressed.

Allele-specific copy number analysis is done by the FACETS v.6.1 [36]. Allele specific HLA loss is determined using LOHHLA as described before [37].

### RNA and TCR ITH scores

Gene- and patient-wise intra-patient heterogeneity scores were calculated using multi-region data. Data was first median-centered to remove any gene-level bias. For each gene, the difference between each pair of samples from the same tumor were calculated. The median difference between the paired-differences was then taken, yielding a gene-specific, patient-specific measure of heterogeneity. This was repeated for all genes, across all tumors, generating a matrix of gene by patient values. Gene intratumor heterogeneity values are summarized as the median value per gene across all tumors in the cohort. Patient intratumor heterogeneity values are summarized as the median value per tumor across all genes. Patient intratumor heterogeneity values represent the expected value of the absolute log2-fold change for a randomly chosen gene within a given tumor.

TCR ITH score is defined as 1 – percentage of shared clonotypes across multiple regions of tumor based on WTS. T cell clones are estimated using MiXCR application on Illumina BaseSpace (http://basespace.illumina.com/apps/). Furthermore, all ITH scores are classified as high versus low using the median as threshold.

### Distinction between dedicated TCRseq and TCR clones inferred from RNAseq using MiXCR

All TCR associated data analysis in this study (including tissue or PBMC) are based on ultra-deep T cell repertoire sequencing (targeted TCRseq) to mitigate undersampling of T cell clones except TCR ITH analysis in Fig. 3b where ITH associated with multiregional sequencing is derived from MiXCR T cell estimates from RNAseq data due to the lack of multiregional TCRseq for all patients.

### ccRCC evolutionary subtypes and intra-tumor DNA Heterogeneity Score

DNA ITH score is calculated as the ratio of subclonal to clonal driver genomic alterations including SNVs, INDELs, and SCNA [14]. A genomic alteration is defined to be subclonal if it is present in less than half of the regions collected in each patient. Patients who enough DNA biopsies are collected are classified into 1 of the 7 ccRCC evolutionary subtypes as described before [14]. We used neighbor joining tree construction in ape package in R [38] for reconstruction of tumor clones. TCGA ITH score was obtained from a previous study as measured by the number of clones estimated per sample using PhyloWGS [39]. Briefly, PhyloWGS is a method to infer tumor evolution evolutionary using the relationships between tumor subpopulations based on variant allele frequencies while considering copy number alterations.

### HLA and TCR diversity

Shannon entropy is calculated to define TCR diversity [40]. We used MiXCR application on Illumina BaseSpace (http://basespace.illumina.com/apps/) for alignment and T cell clonotype identification. Immunarch (https://immunarch.com/) [41] was used for downstream analysis including visualization and data analysis. Morisita index [42] was used to measure clonotype overlap. HLA diversity index is measured as adopted from [43] as described in [25].

### Neoantigen Depletion

The fraction of neoantigens depleted is defined for each sample where pretreatment data was available. We first calculated the neoantigen depletion as the number of neoantigens that were undetectable after therapy but were detected pretreatment. The fraction of neoantigens depleted was then defined as the ratio of the total number of depleted neoantigens over total pretreatment neoantigens. To distinguish neoantigen depletion due to contraction (immune elimination) from evasion, we exclude any neoantigens that were depleted without the presence of HLA LOH (defects in antigen presentation machinery), or reduced expression i.e., log2(FC)< - 1 where FC is the fold change defined as the ratio of post treatment TPM over pretreatment TPM after correction for tumor purity. Conversely, a neoantigen is annotated as deleted due to immune elimination if log2(FC)>=0 and no HLA LOH was detected. Likewise, HERV editing is defined as the median change in the expression of immunogenic HERVs compared to pre-treatment expression. Immunogenic HERVs refers to HERV loci whose expression strongly correlates with TIL abundance, FDR<0.05.

### Weighted Gene Co-expression Network Analysis (WGCNA) and gene signature extraction

We performed WGCNA [44] on all samples where the fraction of neoantigens depleted was available similar to previously described [10]. Briefly, genes with low expression values and invariant genes, that is, genes that were expressed in <5% of samples or had s.d. ≤ 1 for expression (log2 TPM) were filtered together with non-coding genes. The soft power of 6 was chosen based on goodness of fit to a scale-free network. We first annotate modules as JAVELIN or angiogenesis according to the Spearman correlation between the module eigengene and JAVELIN or angiogenesis ssGSEA scores (highest correlation is classified as JAVELIN or angiogenesis module). Likewise, among all modules, the module with the highest Spearman correlation with the fraction of neoantigens depleted was annotated as immune escape module (85 genes). This 85 genes gene signature was strongly associated with PFS of Avelumab plus Axitinib in JAVELIN Renal 101 (HR=1.45, P=0.02, Extended Data Fig. 11a). To further refine this gene signature, we first sorted genes based on their pairwise spearman correlation (Extended Data Fig. 11b) and then selected genes with the highest spearman correlation such that no genes have a spearman correlation <0.6 (Extended Data Fig. 11c). This reduced the number of genes to total of 12 highly correlated genes known as immune escape signature (TIMP1, PXDN, COL15A1, OLFML2B, COL5A2, DLX5, SOX11, KLHDC8A, UNC5A, ADAMTS14, MMP11, FN1). Several genes (ADAMTS14, MMP11, FN1, COL5A1, COL5A2 and TIMP1) in this signature has previously been described as TGF-β-associated extracellular matrix genes that are linked to immune evasion and immunotherapy failure [45].

### Statistical Analysis

All statistical tests were performed in R. To calculate correlations, cor.test with Spearman’s method was used. Tests comparing distributions were performed using wilcox.test. All statistical analyses were two-sided and p-values were Benjamini-Hochberg corrected.

## Results

### The landscape of microenvironmental ITH in ccRCC

To study ITH in ccRCC, we completed ultra-deep (median coverage of 360X) multi-regional whole-exome sequencing and whole-transcriptome sequencing across 142 tumor regions from 29 patients, including 6 untreated and 23 post ICI (see Methods and Table S2). Tumor biopsies were extracted from different regions of the same primary tumor unless specified (Fig. 1a, and b, Table S2). While intra-tumoral genetic heterogeneity in ccRCC is well-described[46], comparatively little is known about the extent of microenvironmental heterogeneity and its relationship to other molecular features of the tumor. To measure the extent of intra-tumoral microenvironmental heterogeneity, we leveraged multi-regional WTS of up to 5 regions from 29 patients. Using single sample gene set enrichment analysis (ssGSEA) of established gene signatures, we quantified the expression of several TME gene expression signatures recently proposed as biomarkers of response to ICIs and anti-angiogenic agents [47] (myeloid signature [8], JAVELIN signature [10], and angiogenesis signature, see Methods and Supplementary S3). We confirmed that these RNA signatures accurately quantified the abundance of key immune populations using matched immunofluorescence data, including statistically significant associations between CD31/angiogenesis (p=0.0003), CD8/JAVELIN T cell signature (p = 0.02), and CD68/Myeloid infiltration (p=0.0013) (Fig. S1).

Microenvironmental signatures demonstrated extensive heterogeneity across tumor regions from the same patient (Fig. 2a). While a small number of patients showed relatively uniform immune infiltration (*e.g.,* NIVO02, Fig. 2a), the significantly more common phenomenon was for patients to exhibit regions both above and below the median score for a microenvironmental feature of interest (*e.g.,* angiogenesis in MR03, JAVELIN/Teffector signatures in NIVO22). Using the myeloid signature (which has previously been associated with poor response to ICI) as an example, we observed most patients cannot be uniquely classified to myeloid enriched or depleted across all tumor regions (Fig. 2c). Given that several of these signatures are under active investigation as biomarkers of response to ICI, we investigated more generally how classification of regions into high/low was affected by ITH. Remarkably, in more than half of the patients, clinically relevant signatures (Angiogenesis, T-effector, Myeloid, and JAVELIN) could not be consistently classified as high or low (Fig. 2b, 2 patients (MR05 and NIVO10) were excluded since WTS data of only one region was available).

**Figure 2.**
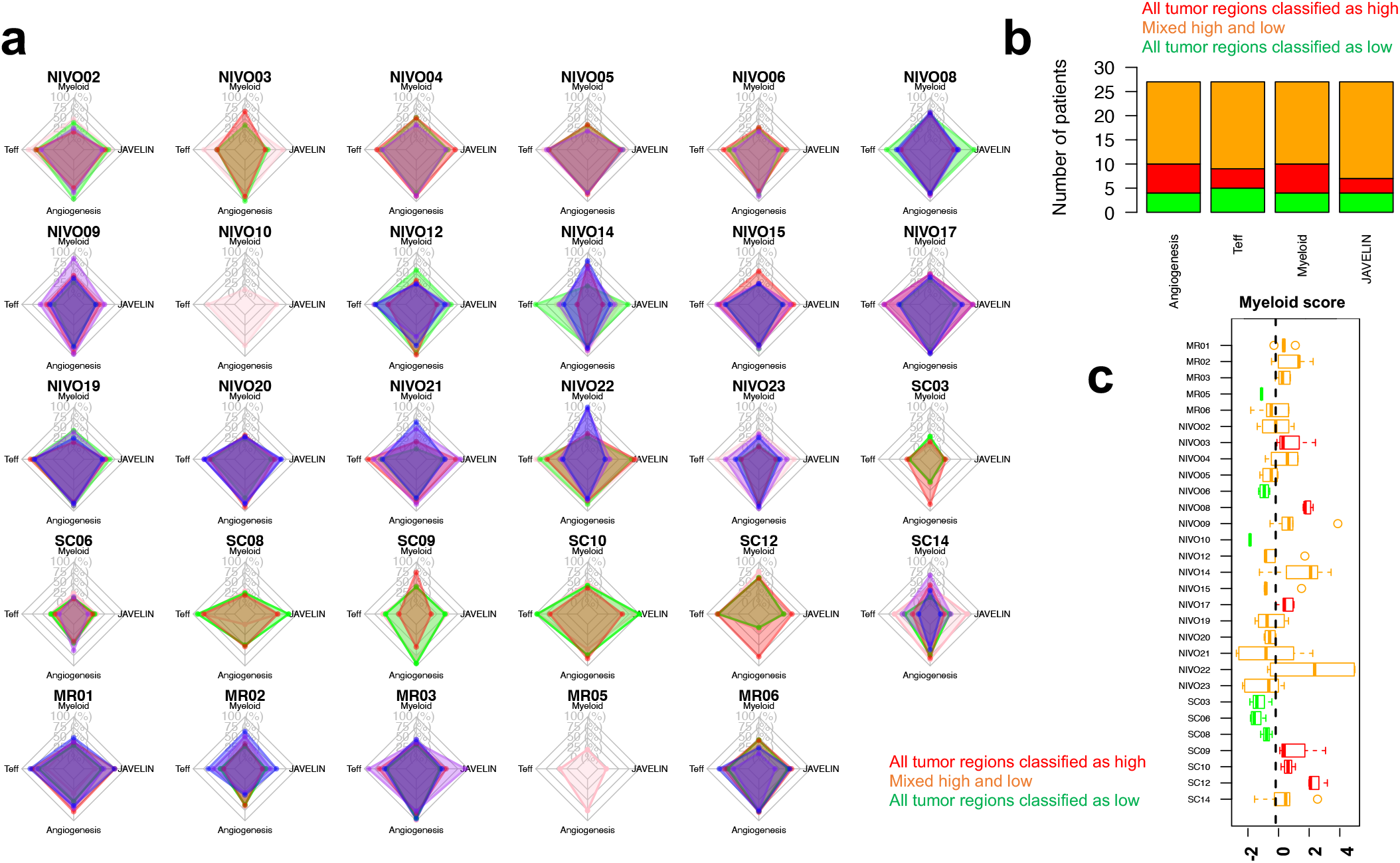
TME ITH in ccRCC. **a)** Intra-tumoral heterogeneity of several gene expression signatures across multiple tumor regions. Radar charts show Z-score of each feature normalized across the cohort. Min and max radius for each feature in each panel represent min and max of that feature across the cohort. **b)** For each gene signature, the number of patients who were classified as high or low or a mixture of high and low across tumor regions are shown. Two patients (MR05 and NIVO10) were excluded since WTS data of only one region was available. Also, pre-treatment regions of ICI treated patients were excluded to avoid treatment-related effects in these signatures. **c)** Intra-tumoral heterogeneity of myeloid score observed across multiple regions of tumors of patients in this study.

We hypothesized elevated microenvironmental heterogeneity may reflect the presence of underlying genomic driver alterations. To test this, we leveraged multi-regional WES data collected for these patients. Frequencies of established ccRCC driver alterations were in agreement with a previous multi-regional study by TRACERx Renal [14] (Fig. 3a). We performed unsupervised hierarchical clustering of major ccRCC driver mutations (i.e., *VHL*, *PBRM1*, *SETD2, BAP1*) and genomic alterations enriched with metastatic disease and ICI response (HLA LOH and CDKN2A/B copy number loss)[37, 43, 48], ultimately identifying two clusters (Fig. 3b, Table S4). We compared the results of these clusters to aggregate, univariate measures of intra-tumoral DNA, RNA, and T-cell receptor (TCR) heterogeneity. Interestingly, one cluster was characterized both by an enrichment of specific genomic alterations (SETD2 mutations, Fisher exact test P=0.002; CDKN2A/B copy number loss, Fisher exact test P=0.0001; HLA LOH, Fisher exact test P=0.0007). This same cluster of patients, which we refer to herein as “ITH-high”, had comparable levels of tumor purity to the other “ITH-low” cluster, but demonstrated elevated ITH at the level of somatic DNA alterations, RNA, and TCR (combined Fisher exact test P=0.0495). Moreover, by classifying patients into previously described ccRCC evolutionary subtypes (Fig. S2), we observed that PBRM1-driven tumors were enriched in the ITH-high cluster (on sample level, Fisher exact test P=0.0018), in agreement with TRACERx Renal [14]. However, this finding must be treated with caution due to our relatively small cohort size as well as low number of regions collected in some patients. These findings were robust to the number of regions collected per tumor, and we found no significant association between ITH and exposure to ICI (Fisher exact test P=0.65, Fig. 3b). Together, our results demonstrate (1) that ITH is not restricted to genomic events, but rather is pervasive in the transcriptome, microenvironment, and immune compartment of ccRCC tumors, and (2) correlates with specific somatic events at the level of individual patients (i.e., PBRM1 and SETD2 mutations, HLA LOH and CDKN2A/B loss).

**Figure 3.**
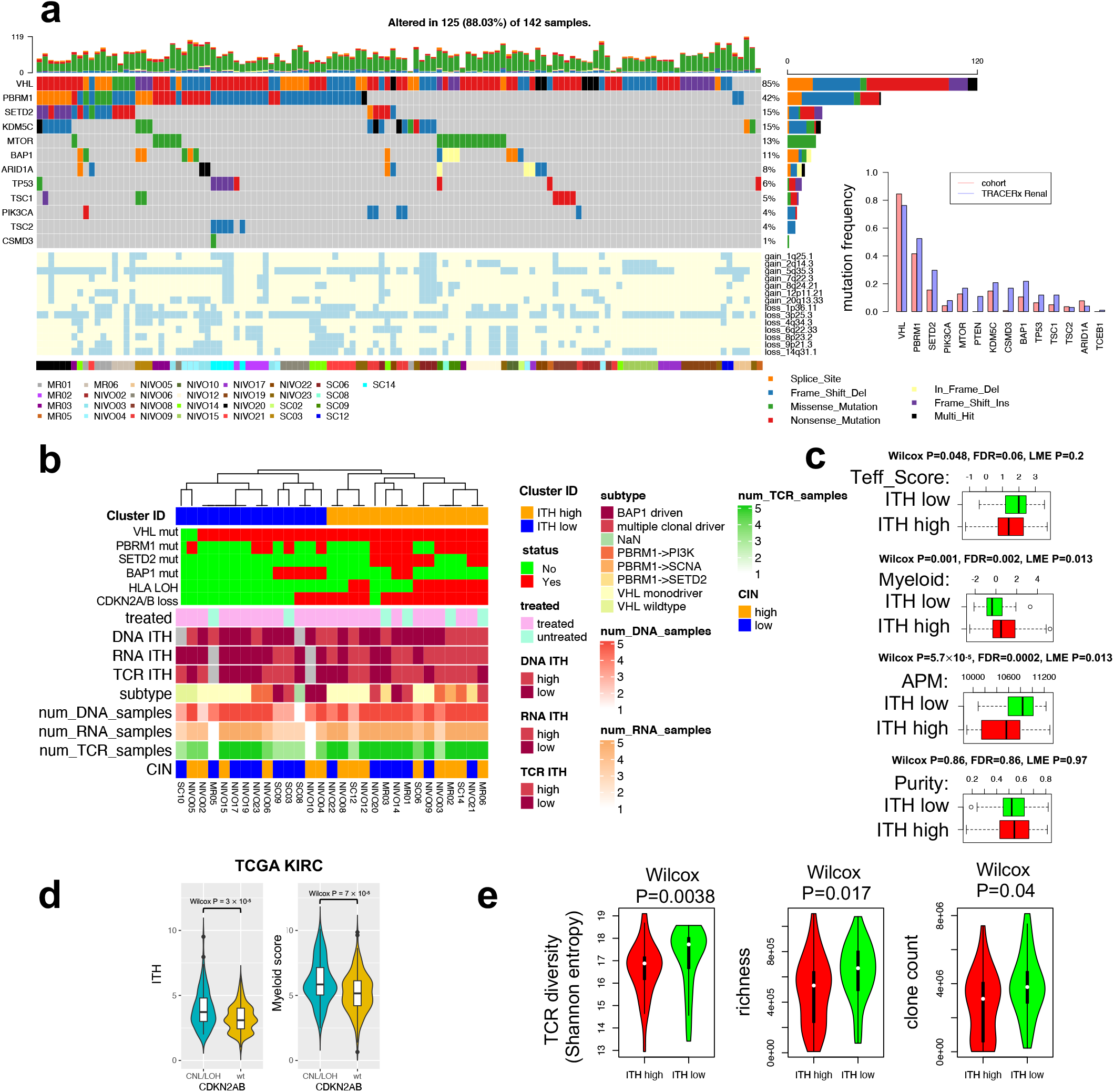
Landscape of ITH in ccRCC. **a)** Oncoprint of key ccRCC driver mutations and copy number alterations for all regions of all 29 patients in this cohort. Margin shows comparison between mutation frequency observed in this cohort and TRACERx Renal. **b)** We have performed unsupervised hierarchical clustering of genomic features including patient level presence or absence of a small variant in VHL, PBRM1, SETD2, BAP1 (most commonly mutated genes) as well as loss of heterozygosity in HLA genes as well as 9p (which includes CDKN2A/B) SCNA which are known to affect ICI response. Heatmap shows ITH high vs low classification across data type. Annotation illustrates evolutionary subtypes and treatment status of patients. A patient is annotated as wildtype if all regions are wild type for that alteration. Cases where ITH score could not be calculated due to lack of sufficient number of biopsies are shown in gray pixels. CIN: chromosome instability. **c)** Association between antigen presentation machinery (APM), effector T cell (Teff) and myeloid gene signatures and ITH. Wilcox P, False Discovery Rate (FDR) and Linear Mixed Effect (LME) P shown. **d)** Intra-tumoral heterogeneity and myeloid score are associated with CDKN2A/B loss in TCGA KIRC cohort. **e)** ITH low patients show a significantly higher TCR diversity, richness and clone count.

### ITH-high ccRCC tumors are immunologically distinct

Comparing the TME characteristics of ITH-high and ITH-low patients, we observed that ITH-high tumors (defined as all regions belonging to a patient who is classified as ITH-high) were characterized by high myeloid and low T cell effector (Teff) signatures (Fig. 3c). Similarly, a signature associated with antigen presentation (APM)[3] was downregulated in ITH-high patients, consistent with elevated levels of HLA LOH in the ITH-high subtype. To validate if genomic features uniquely characterizing ITH-high tumors (HLA LOH and CDKN2A/B loss) might be more generally associated with myeloid infiltration in a large, independent cohort, we obtained DNA and RNA sequencing data from the TCGA KIRC study and scored samples by the presence of CDKN2A/B loss, ITH (as measured by the number of clones estimated per sample using PhyloWGS, see Methods), and myeloid infiltration. This analysis confirmed that in ccRCC, CDKN2A/B loss was associated with higher levels of ITH (P=3×10^-5^) and higher myeloid infiltration (P=7×10^-5^) (Fig. 3d). However, the association between genomic ITH and myeloid infiltration did not reach statistical significance in TCGA KIRC cohort suggesting the association between myeloid infiltration and ITH is likely indirect through certain genomic events such as CDKN2A/B loss.

The findings above suggested that ITH-high tumors may be distinct in their immunophenotype, including in the diversity of their T cell repertoire. We therefore investigated the association between ITH and T cell diversity both peripherally and within the tumor. To do so, we compared the overlap between tissue-resident and peripheral T cells. Repertoire overlap analysis (Fig. S3) illustrated a high degree of shared clonotypes across different tumor regions from the same patient, but a lack of shared clonotypes across patients. ITH-high patients demonstrated a significantly lower peripheral TCR diversity, richness and clone count compared to ITH-low patients (Fig. 3e), suggesting that elevated heterogeneity in the primary tumor is associated with reduced peripheral immunologic diversity in a manner that is consistent with reports in other diseases [49]. To allow a fair comparison of samples with different number of T cells and account for TCR subsampling, we also studied rarefaction curves and estimated TCR diversity by sequentially resampling TCR clonotypes and computing mean number of unique clones [50]. Estimated diversity using rarefaction curves led to a similar conclusion confirming the observed differences in the TCR diversity are unlikely to be due to artifacts in T cell subsampling (Fig. S3b). Together, the above data argue that elevated molecular heterogeneity in ccRCC tumors is associated with a distinct microenvironmental and immunologic phenotype.

### ICI therapy is associated with loss of putative neoantigens and HLA LOH

The clinical management of ccRCC (for which pre-surgical biopsies are often not indicated or used) makes serial profiling of primary tumors on therapy challenging, rendering our understanding of how ICI may remodel tumor physiology incomplete. To overcome this challenge, we took advantage of 16 patients from our neoadjuvant nivolumab clinical trial who had WES performed on their pre-treatment biopsies. This offered a unique opportunity to interrogate both genomic adaptations (including both somatic mutations as well as the expression of potentially immunogenic endogenous retroviral elements, HERVs) to ICI therapy, as well as immunologic changes in the T cell repertoire.

Focusing first on genetic alterations, we anticipated that ICI administration would lead to elimination of some tumor clones and therefore a contraction in total mutation count. However, we observed no consistent trend in the change of either SNV or indel mutational count following ICI therapy (Fig. S4). Nevertheless, the number of non-synonymous SNVs that were predicted to bind to MHC complex *in silico* was consistently reduced across all patients and all biopsies except for NIVO03 (Fig. S4 and Fig. 4a). An opposite trend was observed in the number of putative non-binders, suggesting a selection in favor of non-neoantigenic mutations by tumor during clonal evolution (Fig. 4a).

**Figure 4.**
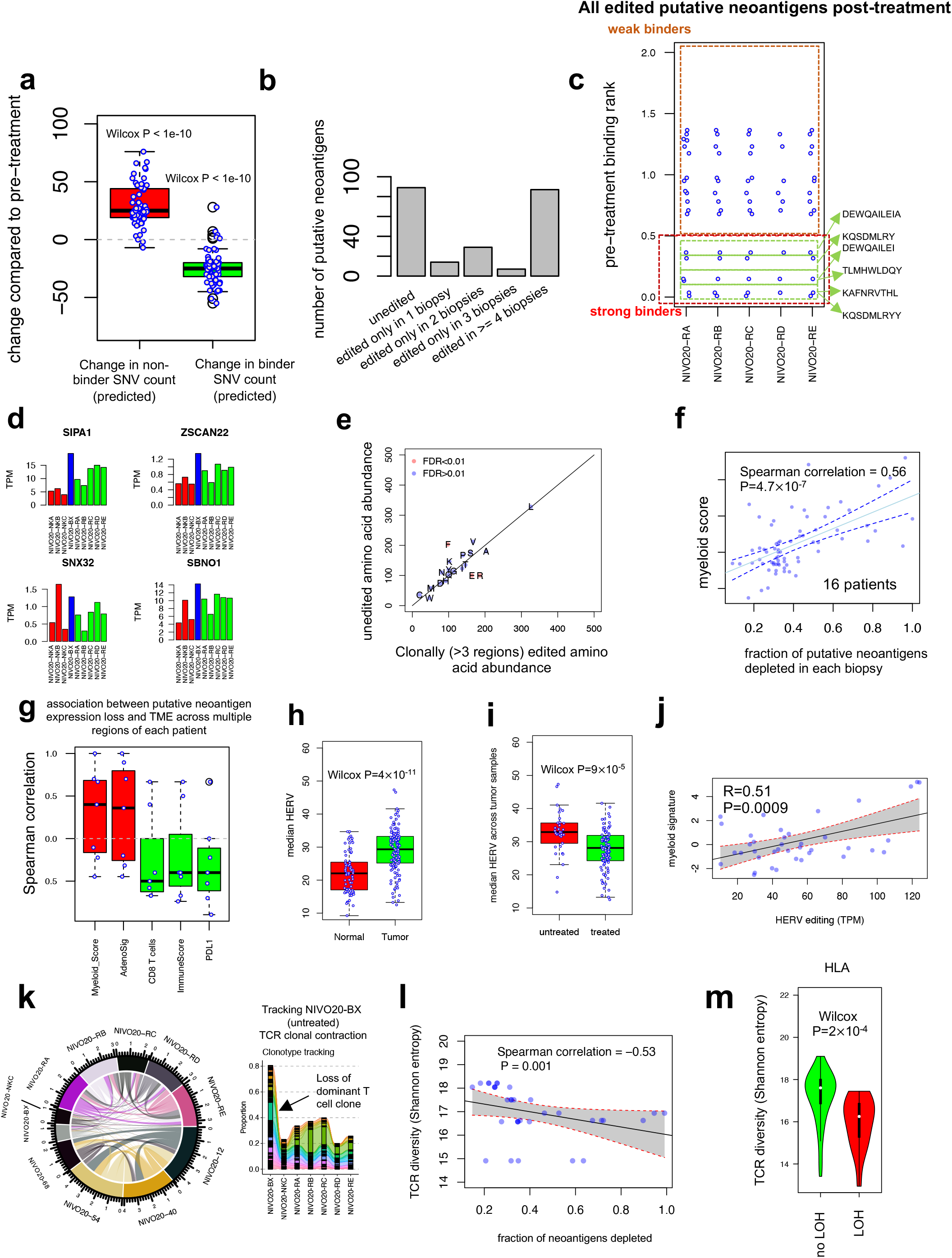
The landscape of heterogeneity of neoantigen depletion. **a)** Change in the number of non-synonymous binder SNVs (predicted *in silico*) and non-synonymous non-binder SNVs compared to pre-treatment. Reduction in only putative neoantigens illustrates selective pressure and immunoediting. One sample Wilcox test P (compared to zero) is shown. **b)** Clonality of neoantigen depletion. Only strong binders are shown. **c, d)** Immunoediting in an HLA-intact patient NIVO20 through reduced neoantigen expression. NKA/NKB/NKC (shown in RED) are normal adjacent tissues 1cm, 2cm, and 4cm away from the center of the tumor; BX (shown in blue) represents pre-treatment biopsy; RA/RB/RC/RD/RE (shown in green) illustrate 5 tumor regions from the treated tumor sample. **e)** Immunoediting with amino acid resolution. Higher Phenylalanine (F) depletion compared to Glutamic Acid (E) and Arginine (R) suggests immune selection. **f)** Association between putative neoantigen depletion and myeloid activation across all regions of patients where pre-treatment WES data was available (n = 16 patients). **g)** Association between the fraction of expressed putative neoantigens depleted and immune signatures. In (**g**) correlations are calculated across different regions of the same patient, for all patients with >3 treated as well as pre-treatment RNA samples were available (n = 7 patients). **h, i)** HERVs are enriched in tumors compared to normal samples and are associated with treatment. **j)** HERV depletion association with myeloid signature. **k)** Circos plot (left) illustrates the fraction of shared T cell clonotypes between tissue and different time points on therapy. Ribbons connecting different regions of the tumor are scaled based on clonotype overlap. (right) clonotype tracking of dominant untreated T cell clones in treated regions of patient NIVO20. The color of each ribbon shows different T cell clones, and the width is scaled corresponding to the frequency of that clone. Tissue data consists of 5 tumor regions after treatment (RA/RB/RC/RD/RE), one single normal adjacent (NKC), and one tumor region pre-treatment (BX). Likewise, PBMC data points on treatment are NIVO20-68, −54, −40, −12. **l, m)** TCR diversity is negatively associated with neoantigen depletion and HLA LOH.

In order to characterize the clonality of putative neoantigen depletion across distinct tumor regions, we counted all 8-11 amino-acid-long putative neoantigens seen prior to treatment but deleted in at least one biopsy after treatment. Among 7 patients with at least 4 tumor regions sequenced, we observed an enrichment for putative neoantigen depletion across 4 or more sites (Fig. 4b). Focusing on patient NIVO20, all 6 identified depleted putative neoantigens were deleted in at least 4 regions, suggesting putative neoantigen depletion is a clonal event (Fig. 4c). Genes expressing these depleted neoantigens demonstrated a 2-3-fold reduction in expression related to pre-treatment biopsy (NIVO20-RA/RB/RC/RD/RE vs NIVO20-BX) (Fig. 4d). Together with the data above, these observations suggest that ICI therapy in ccRCC is associated with the clonal loss of mutations with elevated immunogenicity.

Premised on prior reports [51] of the increased immunogenicity of hydrophobic residues, we sought to determine whether a selective pressure exists on certain neoantigens. We compared the number of amino acids preserved versus depleted upon immunotherapy, and noticed a strong selection against Phenylalanine (F, extremely hydrophobic) in favor of Arginine (R, extremely hydrophilic) and Glutamic acid (E, extremely hydrophilic) in our cohort (Fig. 4e).

We next examined the magnitude of putative neoantigen depletion in each patient by measuring the average number of putative neoantigens deleted per biopsy (i.e., the ratio of the deleted neoantigens in a treated region compared to pre-treatment over the total number of pre-treatment neoantigens). We observed that the fraction of neoantigens depleted was strongly associated with myeloid-high regions (n = 16 patients whose pre-ICI treatment WES data was available, Fig. 4f). The association between myeloid activation and neoantigen depletion remained strong when total number of neoantigens depleted was used (instead of fraction) (Fig. S5b) or when putative neoantigen (transcriptional) expression was taken into account (n = 7 patients whose pre-treatment WTS data was available, Fig. 4g) and was not affected by variation in tumor purity (Fig. S5). Furthermore, the correlation between the degree of neoantigen depletion and myeloid infiltration was also evident when examining different regions of individual patients, where highly depleted regions were associated with the highest myeloid and lowest ImmuneScore (Fig. 4g).

A recent study [52] identified tumor infiltrating lymphocyte specific HERV epitopes that are translated, can bind to MHC I complex, and induce high-avidity cytotoxic T cells. In [52] as well as other previous reports [53], over expression of HERVs on tumor cells has been reported and a link to ICI response has been documented [54]. To interrogate other tumor intrinsic features associated with immune response in our cohort we utilized our deep RNA sequencing (∼200 millions read/library) to quantify HERV expression. HERVs were overexpressed in tumors compared to normal tissues in our cohort (Fig. 4h), and median HERV (median of all HERV loci investigated) was correlated to angiogenic expression (Fig. S7a). Notably, PBRM1 mutations, which lead to further HIF upregulation [55] and angiogenic expression [56, 57], were also positively associated with HERV (Fig. S7b), consistent with a recent report [58]. In agreement with [54] we then confirmed the association between the median expression of different HERV loci and TIL abundance (Fig. S7a). Median HERV was anti-correlated with tumor purity; however, the association between HERV expression and TIL abundance remained valid even when HERV expression was corrected for tumor purity (Fig. S7a). Conversely, we observed a significant reduction in HERV expression an observation akin to reduction in neoantigens (Fig. 4i). Likewise, we observed a strong correlation between HERV editing (i.e., change in the expression of immunogenic HERV loci after treatment, see Methods) and myeloid signature further highlighting the association between neoantigen depletion and myeloid enrichment (Fig. 4j). Due to the limitations of HERV quantification using WTS, we could not rule out that a strong correlation between HERV and TIL abundance might be due to expression of HERV on immune cells. However, the expression of HERV on ccRCC tumor cells has been previously shown [59] and their immunogenicity is well-established [52]. Nevertheless, rigorous determination in future studies of cell-specific expression of HERVs will be critical to understanding their putative association with ICI response.

Finally, using TCRseq of tissue resident and peripheral T cells, we investigated the impact of ICI and neoantigen depletion on T cell diversity. Focusing again on patient NIVO20 where TCR data of multiple regions of pre-treatment and ICI treated tumor were available, we evaluated the degree of overlap between T cell clonotypes at different regions and time points i.e., pre-treatment, on-therapy, and post ICI treatment (Fig. 4k). Tracking dominant tissue resident T cell clonotypes, we noticed a substantial depletion of dominant T cell clones upon ICI therapy (Fig. 4k). This observation was mirrored across our entire cohort, where we observed a strong negative association between peripheral TCR diversity and neoantigen depletion and allele specific HLA loss across the entire cohort where PBMC TCRseq data was collected (Fig. 4l, m). Together, if validated using future mechanistic experiments, our findings suggest that neoantigen depletion in primary ccRCC tumors is associated with peripheral loss of neoantigen reactive T cells. However, at this point, no causal relationship between neoantigen loss and TCR diversity can be drawn.

### Subclonal evolution underlies immune escape

In order to understand the immunologic mechanisms driving subclonal evolution after ICI, we investigated in detail patients whose tumors underwent subclonal immunoediting in distinct regions. Strikingly, subclonal reconstruction revealed recurrent subclonal evolution of HLA LOH and CDKN2A/B loss following ICI therapy (Fig. 5a). Notably, we observed HLA LOH and CDKN2A/B loss co-occur in 9 patients (Fisher exact test P=0.003) and most tumor regions (Fisher exact P=5×10^-7^) (Fig. 5b). Strikingly, comparing the untreated and treated regions, we only observed a significant immunological response (as measured by Th1 response) in regions without CDKN2A/B loss or HLA LOH (Fig. 5c), suggesting that HLALOH or CDKN2A/B loss are subclonal determinants of response to ICI [37, 43, 48]. This is consistent with recently published data [49] indicating the loss of 9p21 - encompassing CDKN2A/B – confers a cold tumor immune microenvironment and resistance to ICI. In that study, Han *et al.* [49] linked 9p21 loss to a decreased abundance of B, T, CD8 T, NK cells and cytotoxic lymphocytes, and an increased fractions of macrophages, as well as reduced TCR CDR3 repertoire abundance and diversity. We interpret our observations to mean that immuno-editing occurs under selective pressure by which certain tumor subclones transform to a less immunogenic phenotype through HLA LOH and CDKN2A/B loss, and that this subclonal selection can produce a highly heterogenous TME.

**Figure 5.**
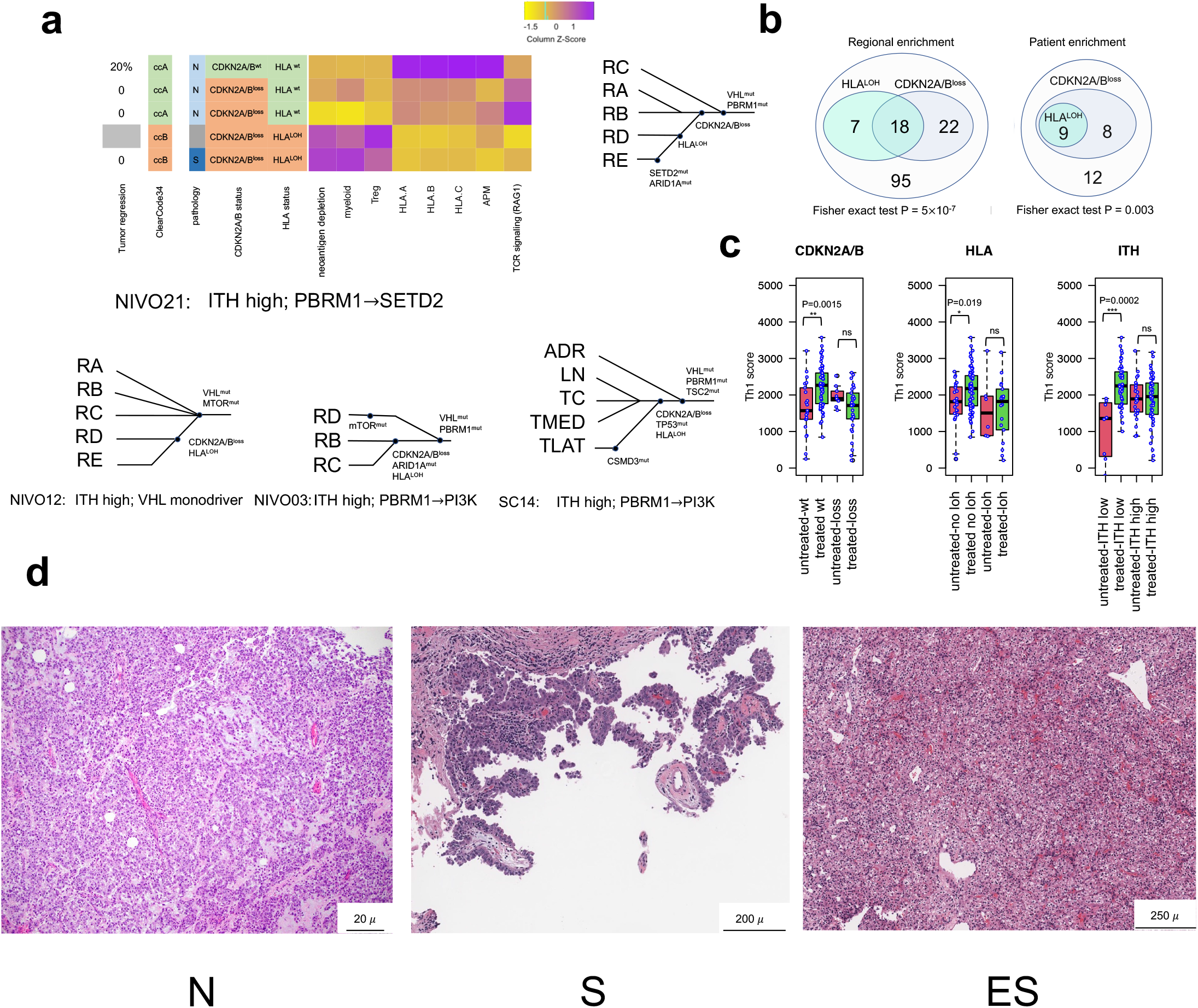
Branch evolution demonstrates immune evasion. **a)** Evolutionary tree illustrates tumors can exploit concurrent HLA LOH and CDKN2A/B loss to escape immune surveillance. **b)** Co-occurrence of HLA LOH and CDKN2A/B can be seen both across regions and patients. **c)** Differential immune response to ICI therapy in patients with CDKN2A/B loss or HLA LOH or belonging to ITH high subtype. **d)** Regions of tumor associated with immune escape depict a distinct pathology where colocalization of TILs and stroma can be observed.

To further shed light on the how tumor evolution can transform TME, we sought to analyze the spatial distribution of TILs within the TME and their interaction with the stromal compartment using immunohistochemical data. Following A.W. Zhang and colleagues [60], a dedicated genitourinary pathologist classified tumor regions into 3 subtypes according to the co-localization of tumor infiltrating lymphocytes and tumor cells: N-TIL (tumors sparsely infiltrated by TILs), S-TIL (tumors dominated by stromal TILs), and ES-TIL (tumors with substantial levels of both epithelial and stromal TILs) (Fig. 5d). We observed that an ES-TIL enriched TME is strongly associated with regions with HLA LOH (ES=4, N=7, S=5 compared to ES=2, N=32, S=21 in HLA intact regions, Fisher’s exact test P = 0.036) or loss of CDKN2A/B (ES=4, N=7, S=9 compared to ES=2, N=32, S=17 in regions without loss of CDKN2A/B, Fisher’s exact test P = 0.03) whereas N-TIL pathology is linked with regions with no HLA LOH and no CDKN2A/B loss across the cohort. These findings suggest that despite abundant TILs, post-ICI ES-TIL are associated with tumor clones that have evolved genetic mechanisms for evasion of the immune response (HLA LOH and/or CDKN2A/B loss). However, future mechanistic studies are needed to pinpoint the primary genomic event that transforms the ccRCC TME into a cold niche.

### An adverse ccRCC TME is enriched stroma and myeloid signatures

We hypothesized that neoantigen depletion could be associated with a specific transcriptional signature, akin to those identified in clinical trial settings as biomarkers for response to ICI in ccRCC. To identify such a signature, we performed unsupervised Weighted Gene Co-expression Network Analysis (WGCNA) [44] to reconstruct modules from our transcriptomic samples similar to [10] (Fig. 6a). Reassuringly, we identified two gene expression modules #7 and #4 reflecting established microenvironmental features associated with therapeutic response in ccRCC: immune inflammatory response (“JAVELIN-like” signature) and “angiogenesis-like” (Fig. 6a, b). We next assessed the correlation between the expression of each WGCNA gene module and neoantigen depletion. While the JAVELIN-like and angiogenesis-like modules showed no association with neoantigen depletion, module 16 demonstrated the strongest association (Fig. 6a). Correlation analysis with previously known gene expression signatures illustrated that module 16 (which we refer to as an “*Immune Escape*” signature) was strongly associated with myeloid and stroma features of TME. The Immune Escape signature also resembled a recently described pan-cancer TGFβ signature derived in a previous study [45] which was linked to cancer-associated fibroblasts enriched in immune evasion and immunotherapy failure. However, no association between the Immune Escape signature and treatment status was observed (Wilcox P=0.79) (Fig. S8).

**Figure 6.**
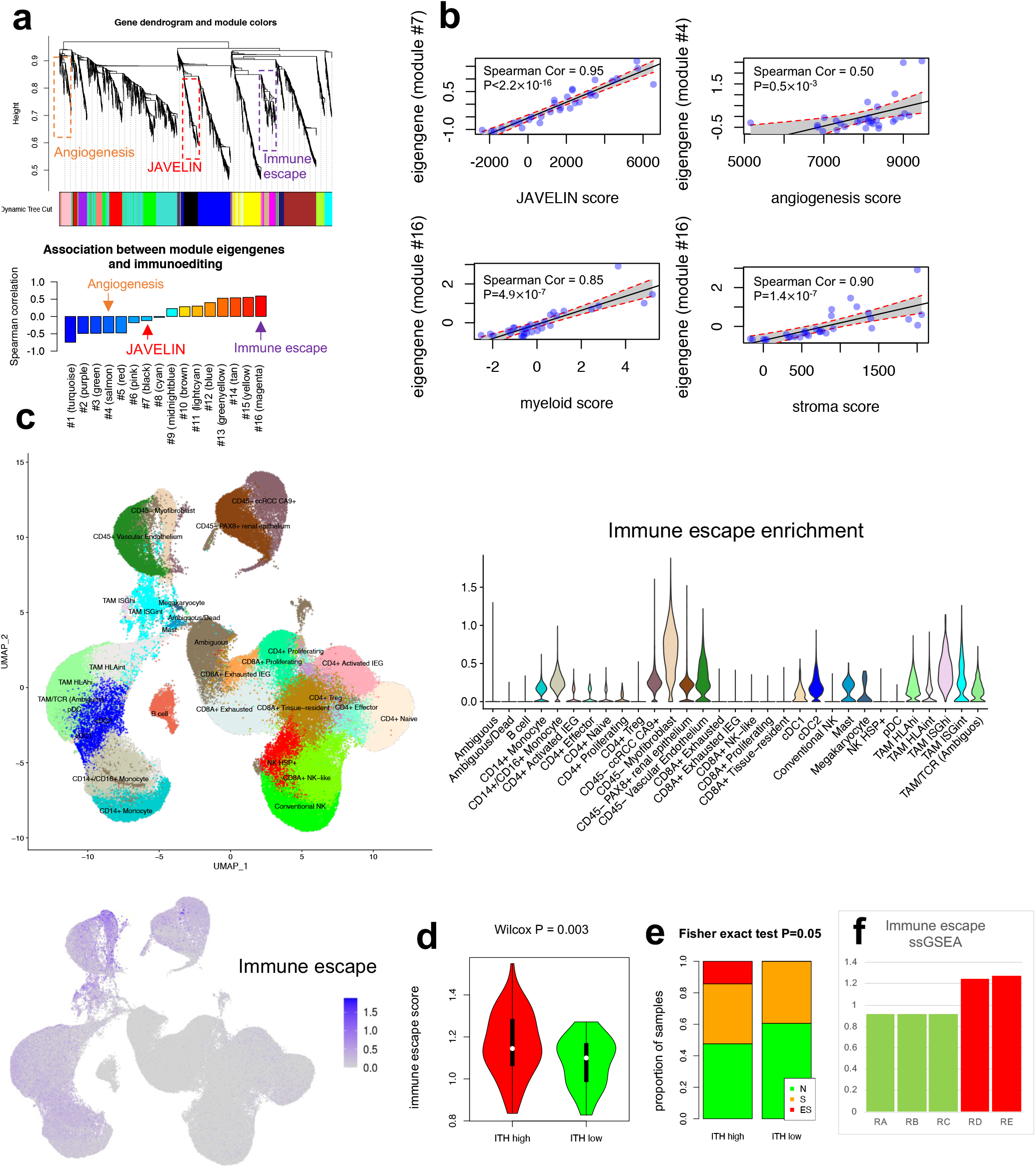
Immunoediting correlates with stroma and myeloid signatures. **a)** WGCNA identifies gene expression modules associated with inflammation (“JAVELIN-like”), angiogenesis, and Immune Escape. Gene dendrogram was first generated and then modules were extracted using dynamic tree cutting (top). Modules were annotated by comparing the correlation between the module eigengenes and previously known gene signatures describing different phenotypes (bottom). **b)** Modules 7 (black), 4 (salmon), and 16 (magenta) are associated with previously described signatures, JAVELIN, angiogenesis and myeloid/stroma. **c)** scRNAseq demonstrates the cell type enrichment of Immune Escape signature in ccRCC patients. Different colors represent different cell types inferred from scRNAseq data. UMAP plot illustrates single cells collected from all 6 patients including treated and untreated patients. Computational extracted clusters were annotated as previously described [15] **d, e)** Association between ITH groups, Immune Escape signature and N/S/ES pathologies. **f)** These regions demonstrate an elevated Immune Escape gene signature in NIVO21. RA, RB, RC, RD, and RE denote different regions of a tumor sample.

To reveal the primary cellular populations driving the Immune Escape signature in the ccRCC TME, we leveraged scRNAseq from multiple tumor regions, lymph node, normal kidney, and peripheral blood of two ICI-naïve and four ICI-treated patients [15] (n=167283 single cells) (Fig. 1c). We identified 28 clusters (Fig. 6c) using Louvain clustering [61, 62] and each cluster was annotated based on our previous study [15]. As expected, scRNAseq revealed enrichment of this signature in renal epithelium, tumor stroma as well as tumor associated macrophages (TAMs) and monocytes (Fig. 6c). Hence, both scRNAseq and histopathological evaluation further confirmed the association between Immune Escape and neoantigen depletion (Fig. 6a, Spearman correlation = 0.6), ITH (Fig. 6d, Wilcox P=0.003), myeloid activation (Fig. 6b, Spearman correlation = 0.8) and with stroma, and renal epithelium histopathology (Fig. 5d and Fig. 6e, f).

### Immune Escape correlates with clinical outcome to ICI therapy

Several previous studies have associated signatures of Immune Escape with poor clinical outcome in ICI treated patients [63]. Thus, we evaluated whether our Immune Escape signature can correlate with clinical outcome to ICI treatment. We obtained publicly available RNAseq data for several clinical trials including phase 3 JAVELIN Renal 101 trial [10] – a phase III randomized anti-PD-L1 (avelumab) plus tyrosine kinase inhibitor (TKI, axitinib) versus multi-target TKI (sunitinib), IMmotion151 [64] – a phase III trial comparing anti-PDL1 (atezolizumab) plus anti-angeniogenesis agent (bevacizumab) versus TKI (sunitinib) in first-line metastatic renal cell carcinoma, CheckMate 009/010 – a phase I/II, aPD-1 (nivolumab) treated, and CheckMate 025 – a phase III randomized mTOR inhibitor (everolimus) versus aPD-1 [9]. We stratified patients by the median score (see Methods) of the 3 gene signatures obtained in our study (i.e., module 4/JAVELIN_like, 7/angiogenesis-like, and 16/immune escape.

The Immune Escape signature was strongly associated with the response to all three ICI regimens (avelumab plus axitinib HR=1.53 P=0.008, atezolizumab plus bevacizumab HR=1.35 P=0.019 and nivolumab HR=1.45 P=0.02, Fig. 7 and Fig. S9). In contrast, the JAVELIN-like inflammatory signature was strongly associated with clinical outcome to avelumab plus axitinib (HR=0.64 P=0.006), but no association with clinical benefit was found between atezolizumab plus bevacizumab (HR=0.82 P=0.126) or nivolumab treatment (HR=0.97 P=0.823) (Fig. 7). Similarly, the angiogenesis-like signature was strongly correlated with the response to sunitinib in both IMmotion151 (HR=0.48 P<0.001) and JAVELIN Renal 101 (HR=0.68 P=0.008) as expected, but not with ICI-associated regimens. Associations between the Immune Escape signature and therapeutic response remained valid even when thresholds other than median were used to define immune escape high and low (Fig. S10). Even though the Immune Escape signature was also associated with response to sunitinib in JAVELIN Renal 101, no association between sunitinib response or mTOR inhibition was observed in IMmotion151 and CheckMate 025. Overall, this analysis suggests that a transcriptional signature associated the tendency to lose putative neoantigens after ICI is associated with response to combination ICI therapy and nominates a new potential biomarker for this therapeutic regimen.

**Figure 7.**
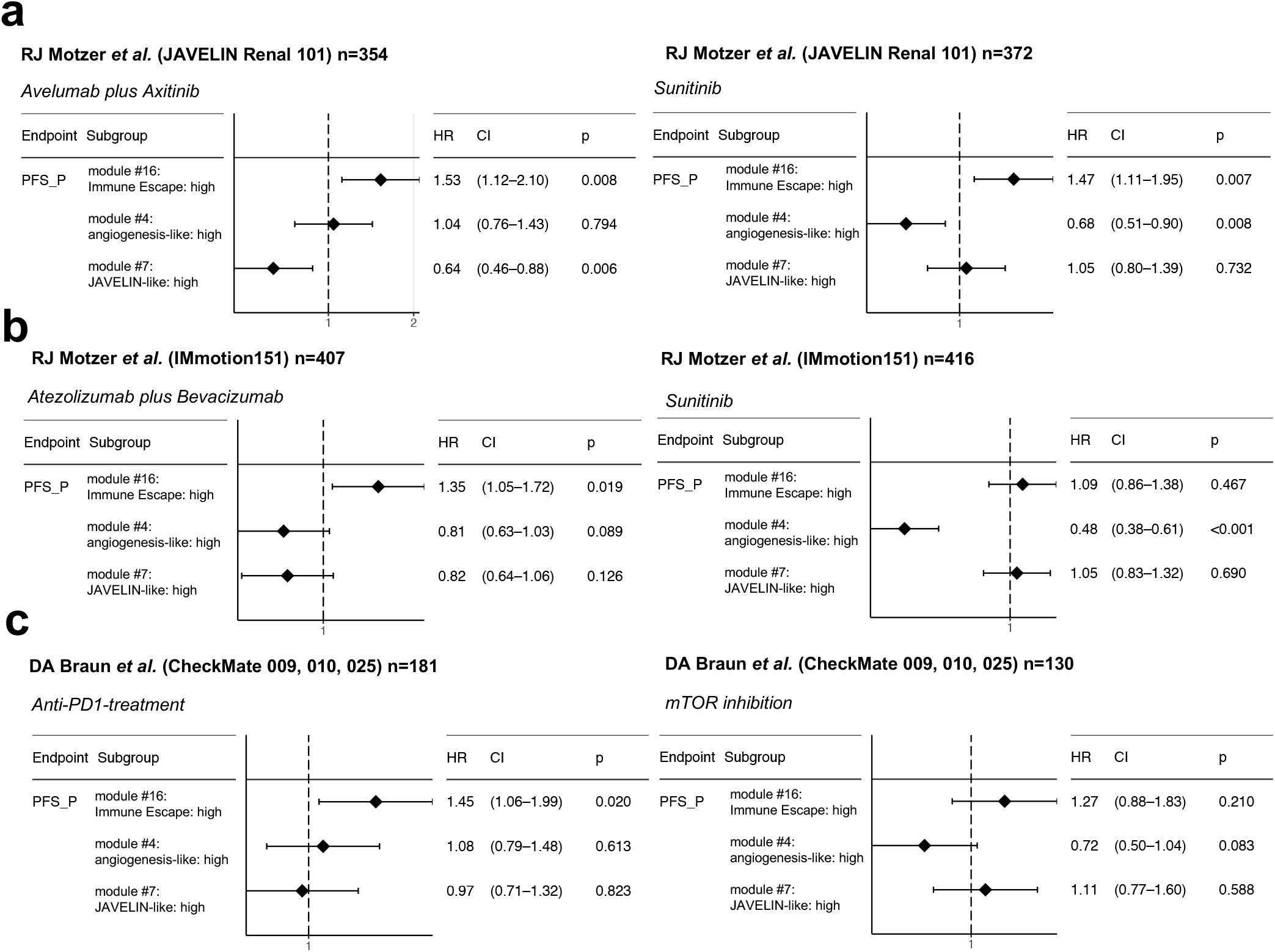
Association between immune escape and clinical outcome to checkpoint blockade. Survival analysis shows the association between gene signatures obtained in this study and clinical outcome of different independent retrospective trials. **a)** Immune Escape and JAVELIN-like signatures are associated with PFS in patients treated Avelumab plus Axitinib in JAVELIN Renal 101 cohort. **b)** Immune Escape signature, but not the JAVELIN-like signature, correlates with the response to Atezolizumab plus Bevacizumab in IMmotion151 but not JAVELIN signature. **c)** Immune Escape signature, but not the JAVELIN-like signature, correlates with the efficacy of anti-PD1-treament in CheckMate 009, 010, 025.

## Discussion

Here we used spatiotemporal, multimodal profiling to investigate the link tumor genomics, microenvironmental heterogeneity, peripheral immune response, and eventual immune escape in advanced and metastatic ccRCC. The fundamental discovery of our analysis is that ITH manifests well beyond the tumor genome and produces highly heterogeneous immune microenvironments in the tumor. Our findings clearly suggest that the ccRCC genome and microenvironment co-evolve, and that loss of putative neo-antigens (including SNVs, indels, and HERVs) is associated with a qualitatively myeloid-high environment and the loss of HLA and CDKN2A/B. These distinct genomic alterations are also associated with more peripheral changes, i.e., reduced T cell clonal diversity in the peripheral circulation.

Emerging data on biomarkers of response to ICI in ccRCC has identified two potentially paradoxical observations: first, that TIL abundance alone is an insufficient predictor of ICI response [9], and second, that the presence of myeloid cells correlate with resistance to both ICI and anti-VEGF treatments. Strikingly, we observed that high myeloid score tumors were associated with neoantigen depletion which could, in principle, render ICI treatment ineffective. In agreement with this, we derived a transcriptomic signature associated with neoantigen depletion and Immune Escape, which was expressed in renal epithelium, tumor stroma as well as tumor associated macrophages (TAMs) and monocytes. This Immune Escape signature was associated with response to several ICI regiments in published clinical trials. In total, these findings suggest that myeloid cells are associated with tumor clones that have evolved mechanisms to escape anti-tumor immune responses. Critically, such a hypothetic model requires detailed work and mechanistic validation in immunocompetent systems, which we are actively developing.

Why do regions with neoantigen depletion demonstrate elevation of myeloid cells but not cytotoxic T cells that would presumably eliminate tumor clones? Cancer immunoediting proceeds through three phases: elimination, equilibrium and escape [65]. Throughout these phases, tumor immunogenicity evolves, and thereby, despite possible initial response to therapy, acquires immunosuppressive mechanisms that may enable disease progression. Our data suggests that myeloid-high, neoantigen-depleted tumor regions historically experienced a cytotoxic T cell response, which prompted the selection of tumor clones losing neoantigens and/or HLA/CDKN2A/B. Such a loss of target antigens through HLA LOH or neoantigen depletion would result in loss of antigen-TCR stimulation, leading to death of the corresponding neoantigen reactive T cells (Fig. 8). Importantly, as with other findings in this analysis, the association between neoantigen loss and myeloid activation observed in our data remains purely correlative, and future studies will be necessary to mechanistically establish how immune evasion spatiotemporally evolves in ccRCC following ICI therapy.

**Figure 8.**
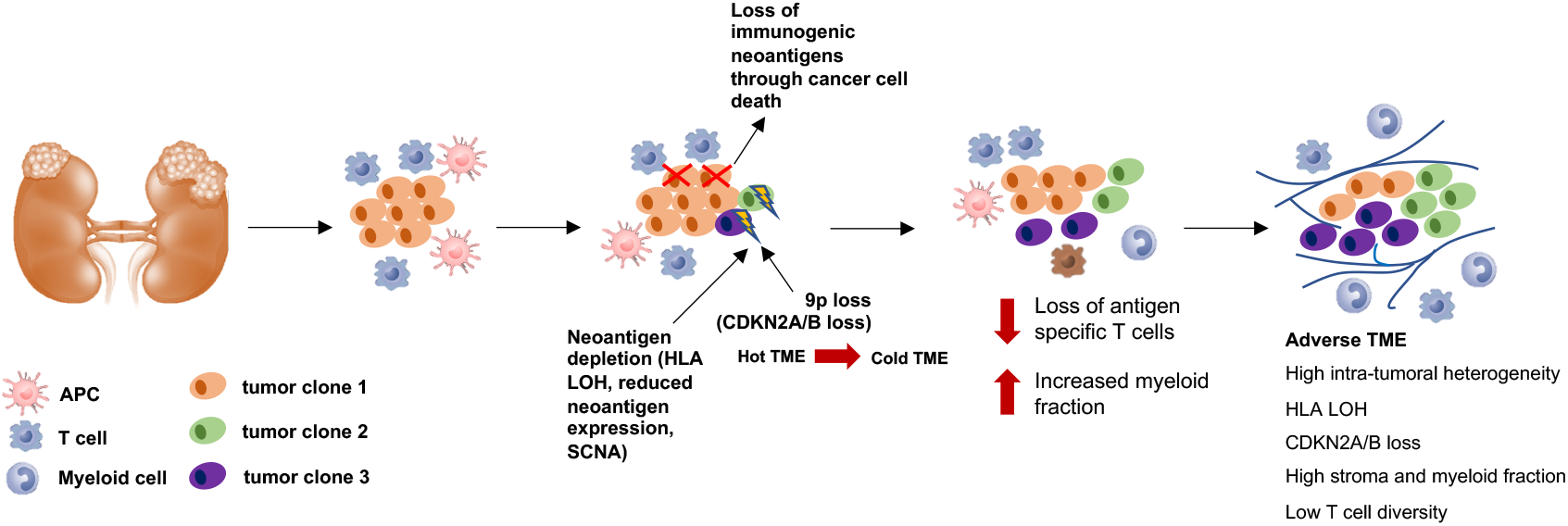
A hypothetical model for spatiotemporal evolution of ccRCC links ITH to immune escape and adverse TME. Cancer cell death, potentially by cytotoxic killing, early in tumor evolution selects for tumor clones with HLA LOH and/or CDKN2A/B loss. This promotes the evolution of a TME depleted of antigen-specific T cells and enriched for myeloid cells.

Our multi-regional data also has significant implications for biomarker development. We demonstrated that TME markers of response such as JAVELIN and myeloid scores can be heterogenous within tumors regions (Fig. 2). This underscores the importance of accounting for ITH when these signatures are used for patient selection for a specific therapy and longitudinal monitoring of therapies. Given recent data that ICI may have a role in adjuvant therapy following nephrectomy for high-risk disease, our data would suggest that several regions of the primary tumor should be sampled specially in the presence of ITH associated genomic alterations (e.g., HLA LOH and CDKN2A/B loss). An intriguing finding was a trend towards lower ITH in ICI treated tumors, even though this observation did not reach statistical significance. If validated in other studies, this in part can be attributed to outgrowth of few non-immunogenic tumor subclones that managed to escape immune surveillance upon ICI treatment.

An important limitation of this study is that TME heterogeneity of metastatic disease was not assessed and may be less of an issue in biomarker development. Our study has several other potential limitations including its small sample size. To overcome this shortcoming, we validated several of our major findings in several independent cohorts. Another potential limitation of our study is the unavailability pre-treatment multi-regional sequencing data. However, inclusion of multi-regional data from 6 untreated patients allowed us to account for ITH in untreated tumors. Moreover, our neoadjuvant cohort was treated with single agent nivolumab over a short course which may not reflect the TME, and genomic changes induced by more potent combination strategies. Finally, we portrayed the characteristics of an adverse TME which may contribute to ICI resistance. Our study clearly demonstrates the interplay between genomic events and TME transformation from a cytotoxic to a cold immuno-phenotype. However, these findings remain purely an association of several contributing factors to ICI resistant and the exact causative hierarchy of events requires further investigation.

In conclusion, we find distinct genomic event enriched in immune escape tumor microenvironment in ccRCC both across and within tumors. Our findings have implications for future biomarker development for ICI response across ccRCC and other solid tumors.

## Data Availability

All data produced in the present study are available upon reasonable request to the authors

## Availability of data and materials

No new code was generated. All data generated in this study are provided in Extended Data Table and Figures or available upon reasonable request from corresponding author.

## Abbreviations

TCGA: The Cancer Genome Atlas
WES: Whole-exome sequencing
WTS: Whole-transcriptome sequencing
SCNA: Somatic copy number alterations
LOH: Loss of heterozygosity
ITH: Intra-tumoral heterogeneity
TCR: T cell receptor
ccRCC: clear cell Renal Cell Carcinoma
TME: Tumor microenvironment
ICI: Immune checkpoint inhibitor
TKI: Tyrosine kinase inhibitor
HERV: Human endogenous retrovirus
PBMC: peripheral blood mononuclear cells
TIL: Tumor infiltrating lymphocytes
WGCNA: Weighted gene co-expression network analysis
ssGSEA: single sample gene set enrichment analysis

## Contributions

M.G. performed data analysis and wrote the manuscript with input from all authors. M.G., A.A.H., T.A.C., and E.R. conceived the study and contributed to data interpretation. F.K., E.R., C.T., V.M., S.Z., C.Z., and R.M. assisted with analytical methodology development and bioinformatics support. M.L.S., R.V., L.L., T.P., and J.G. contributed to DNA, RNA, and TCR sequencing. R.J.M., M.I.C., P.R., J.C., L.G.T.M., and M.H.V. cared for patients analyzed in the study. K.A.B. and R.G.N. handled patient samples. R.G.N. collected and analyzed clinical metadata. S.G. and Y.C. analyzed H&E images. All authors read and approved the manuscript.

## Ethics declarations

### Ethics approval and consent to participate

Informed consent and institutional review board approval were acquired at Memorial Sloan Kettering Cancer Center (MSK).

### Consent for publication

Not applicable.

## Supplementary Figure Legends

**Fig. S1.**
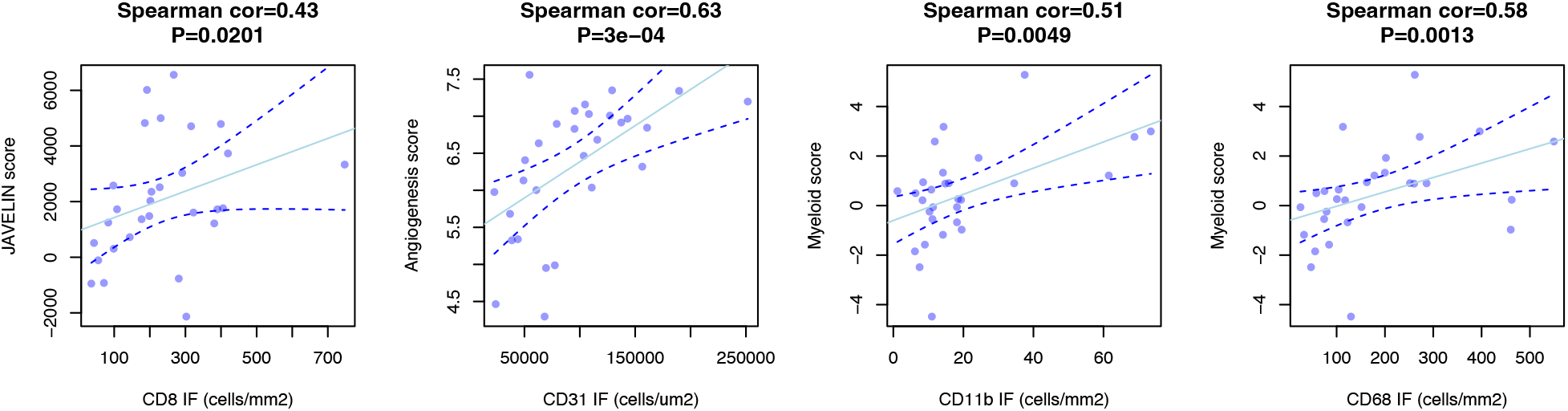
Validation of TME associated gene signatures using IF. Myeloid signature correlates with CD11b/CD68 markers. CD31 endothelial and CD8 T cell markers are correlated with Angiogenesis and JAVELIN signatures.

**Fig. S2.**
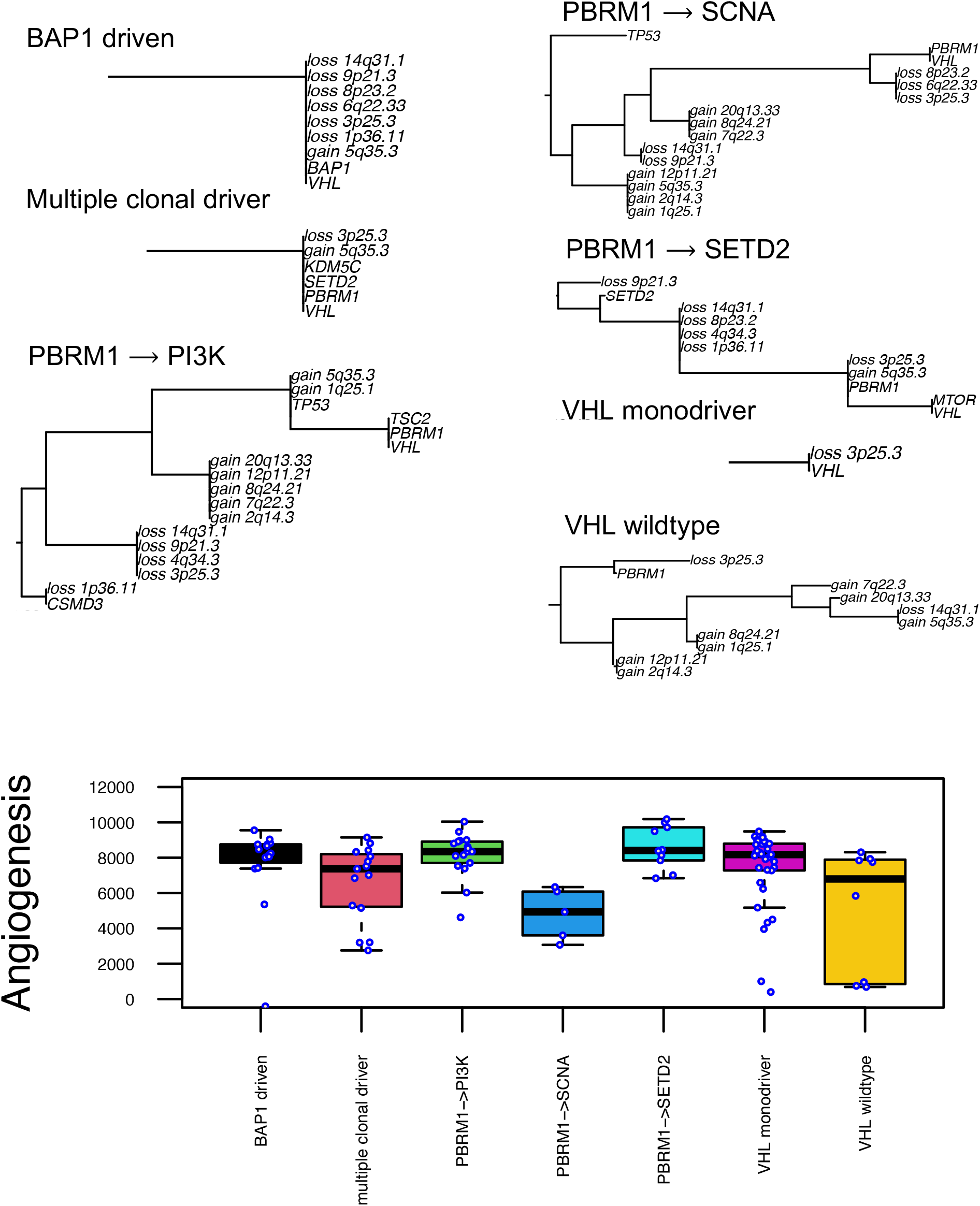
ccRCC evolutionary subtypes and their association with angiogenic TME score.

**Fig. S3.**
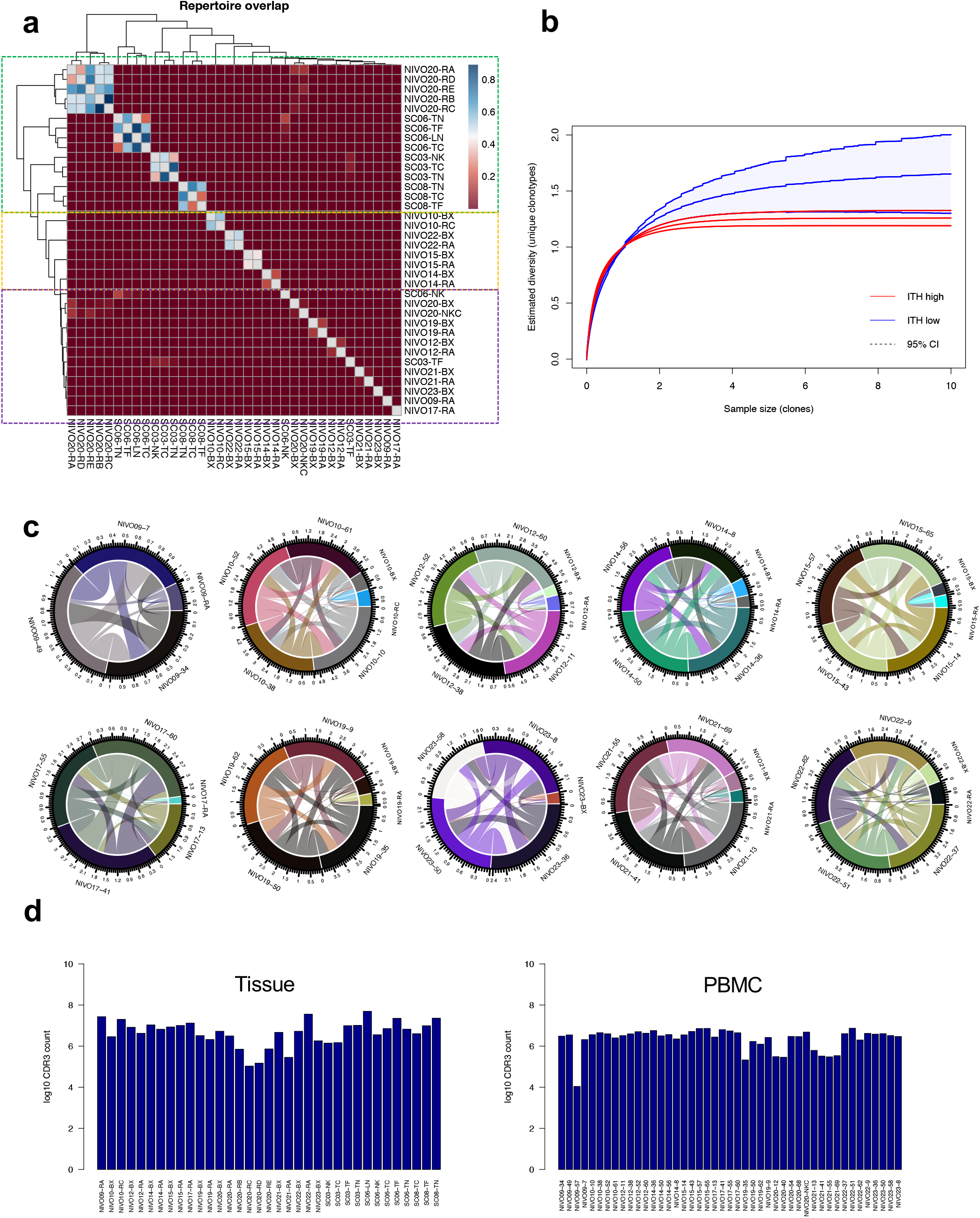
Clonotype overlap analysis. **a)** Hierarchical clustering of TCR clonotypes across different regions of patients where tissue TCRseq data was available. **B)** Rarefration analysis of ITH high vs ITH low patients **c)** Circos plot illustrates the fraction of shared T cell clonotypes between tissue and different time points on therapy. Ribbons connecting different regions of the tumor are scaled based on clonotype overlap. Labels show patient ID followed by time to nephrectomy (e.g., NIVO09-7: patient NIVO09 at 7 days to nephrectomy) **d)** Number of CDR3 clone counts shown for all samples where TCR seq was performed.

**Fig. S4.**
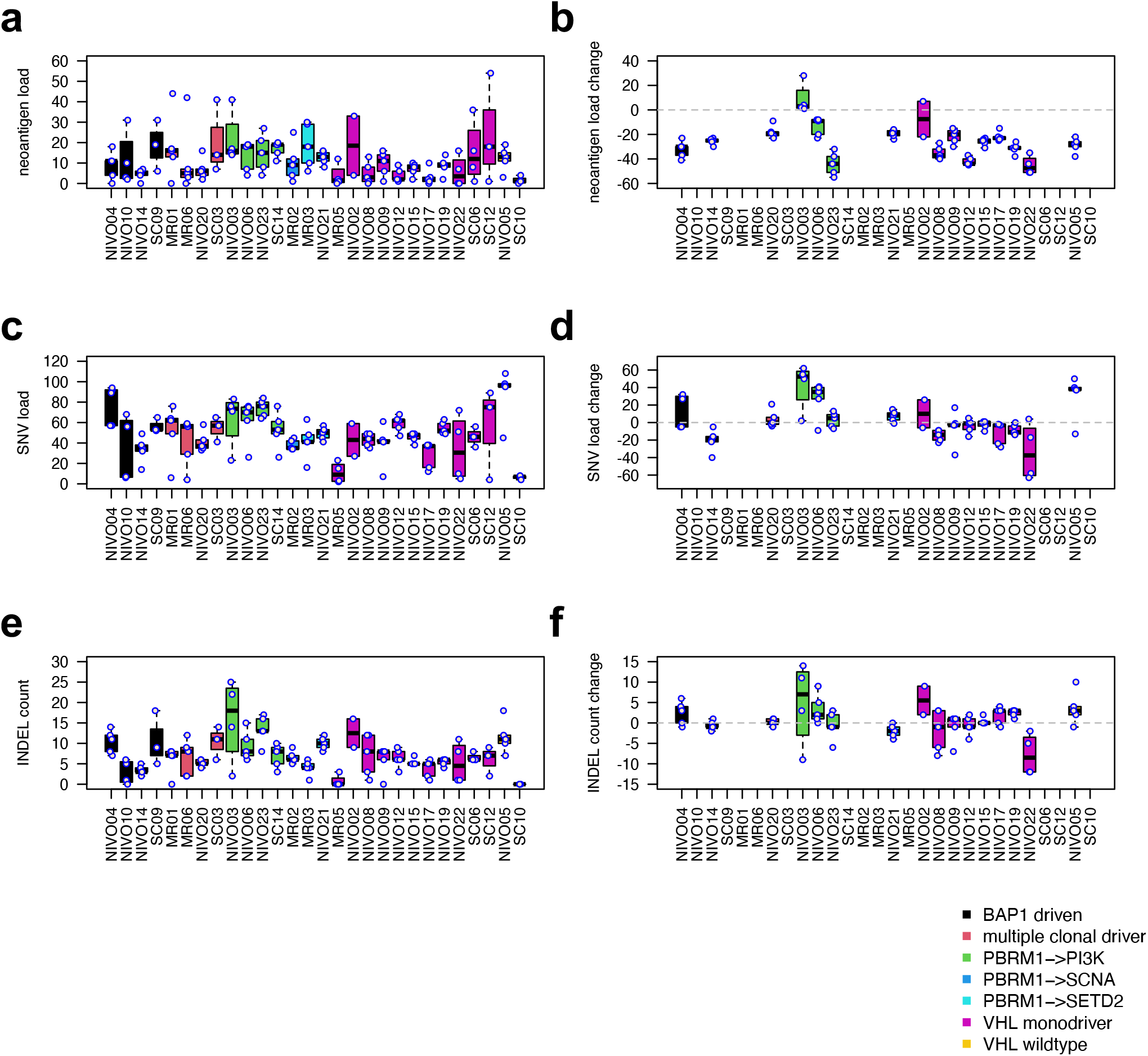
Boxplots show total and change compared to pre-treatment (when sample was available) for mutational count, and neoantigen count across different regions of all patients. Count change is shown only for 16 patients whose pre-treatment WES data was available.

**Fig. S5.**
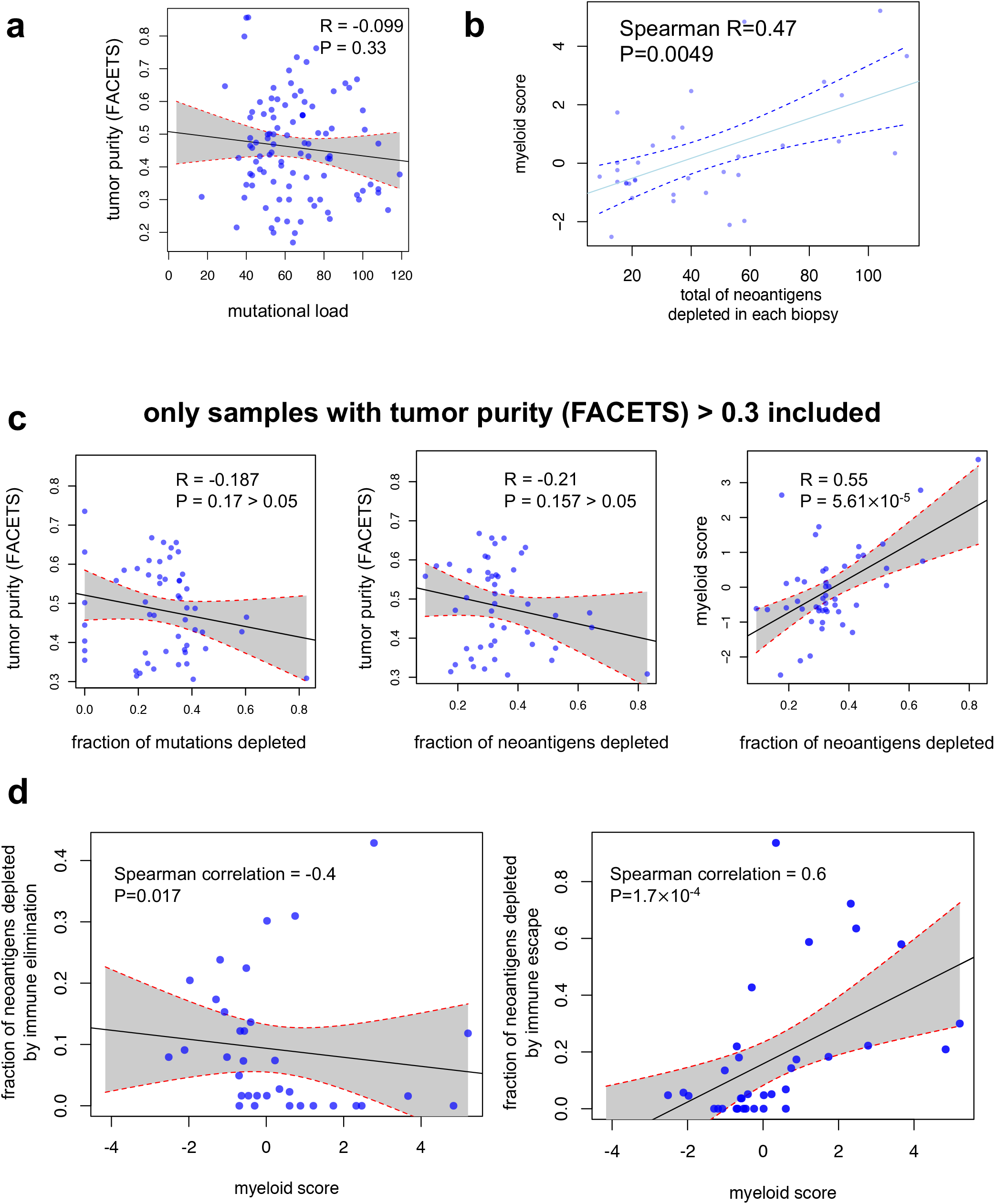
Association between neoantigen loss and myeloid signature. **a)** No association between TMB and tumor purity (FACETS) was observed **b)** Association between total number of neoantigens depleted and myeloid signature. **c)** The fraction of mutations or neoantigens depleted is not correlated with tumor purity for samples with tumor purity larger than 0.3; however, the association between myeloid signature and neoantigen depletion remains strong even after excluding samples with low purity. **d)** Association between immune elimination and escape with myeloid signature.

**Fig. S6.**
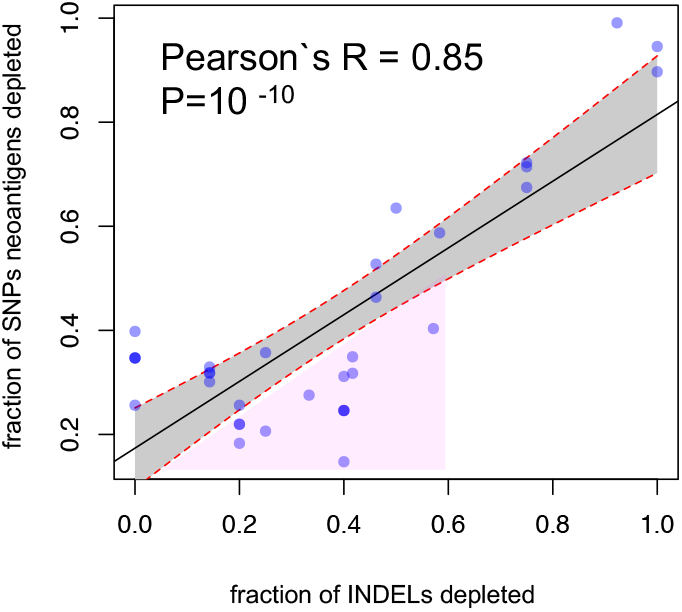
Comparison between SNPs neoantigen depletion and INDEL depletion.

**Fig. S7.**
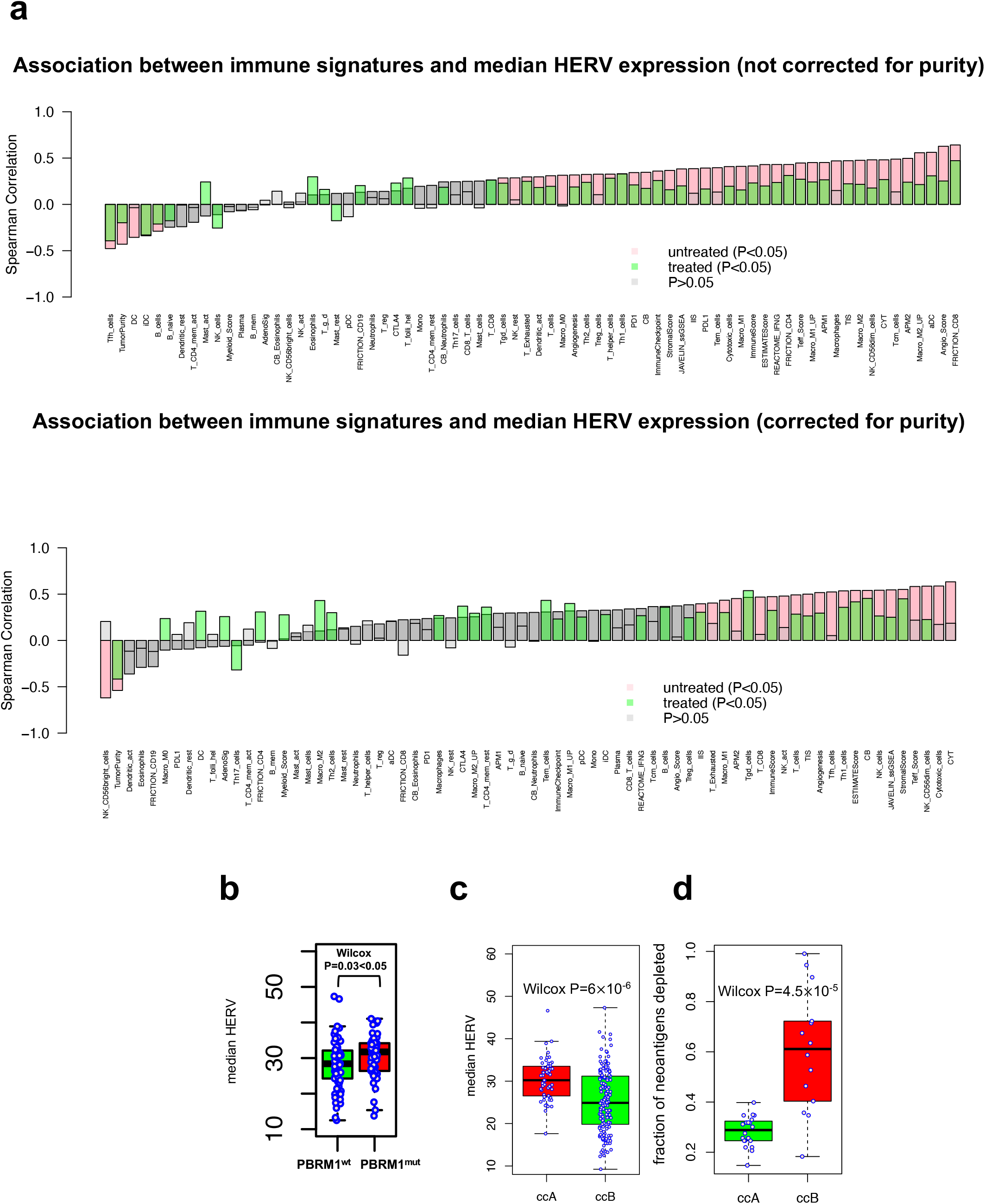
Treatment impact of HERV association with TME. **a)** Association between HERV expression and immune signatures. **b)** PBRM1 mutations are associated with elevated HERV expression**. c)** Association between ClearCode34 classes and HERV expression. **d)** Association between ClearCode34 classes and neoantigen depletion.

**Fig. S8.**
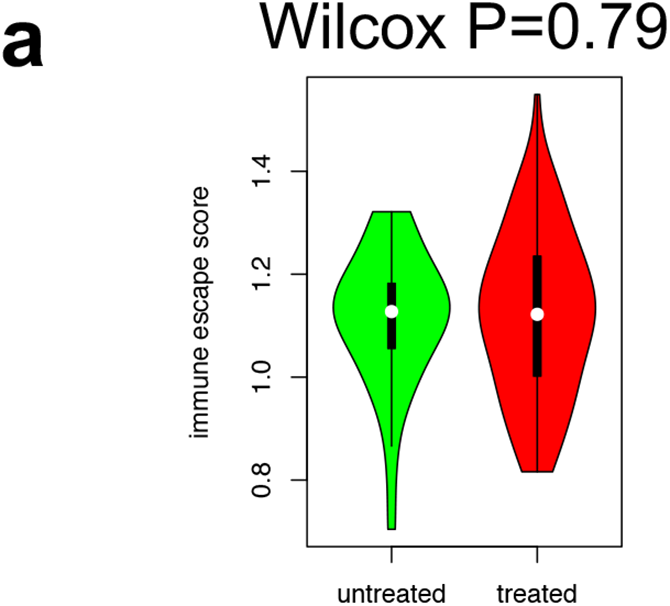
Association between immune escape signature and treatment.

**Fig. S9.**
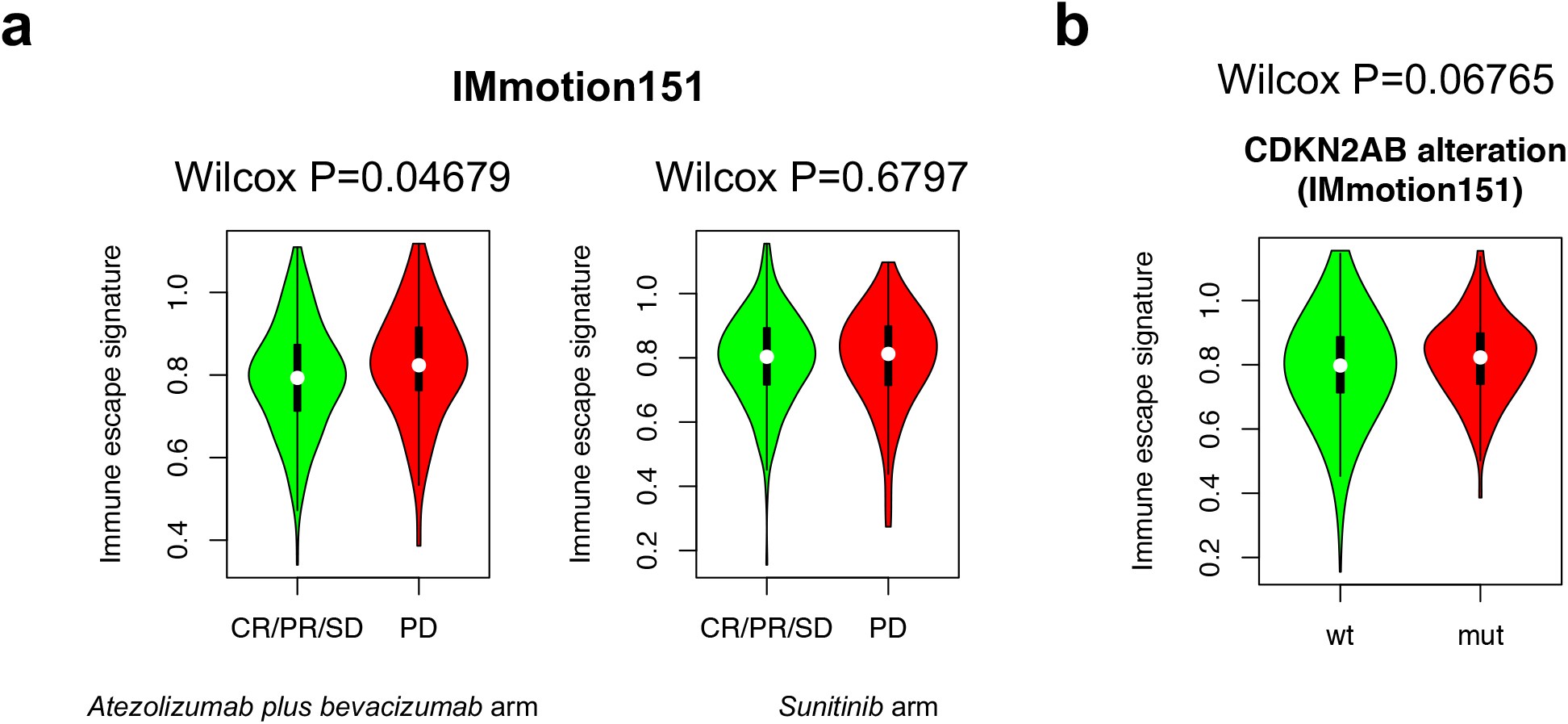
Validation of escape signature in independent cohorts (IMmotion151). **a)** Escape signature is associated with improved survival in patients treated with ICI but not sunitinib**. b)** Escape signature is associated with CDKN2A/B alteration in Immotion151.

**Fig. S10.**
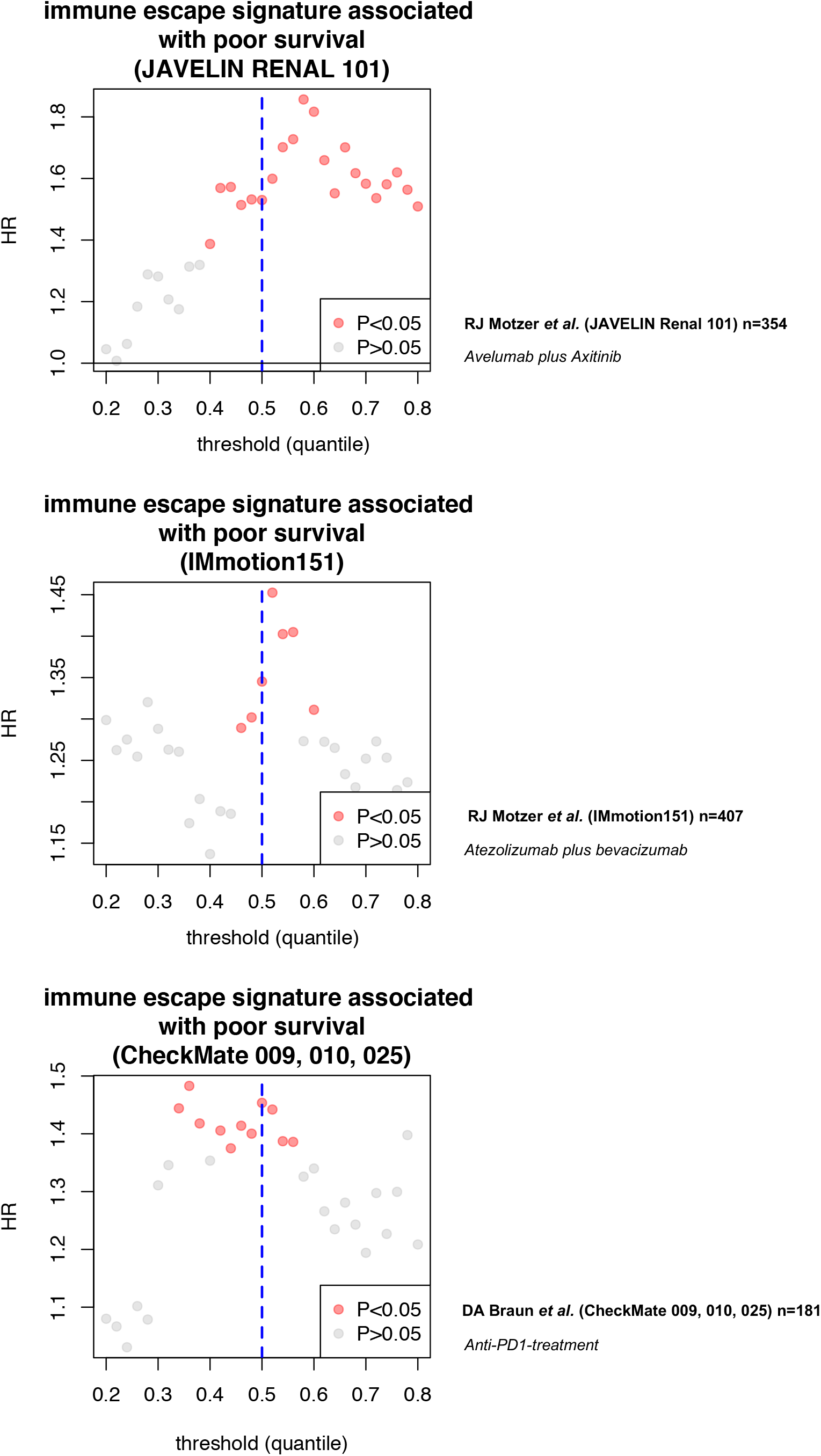
Relationship between escape gene signature and treatment outcome in different clinical trials. HRs are calculated for each threshold for ICI or ICI in combination with TKI arms in JAVELIN Renal 101, IMmotion151, and CheckMate 009, 010, 025.

**Fig. S11.**
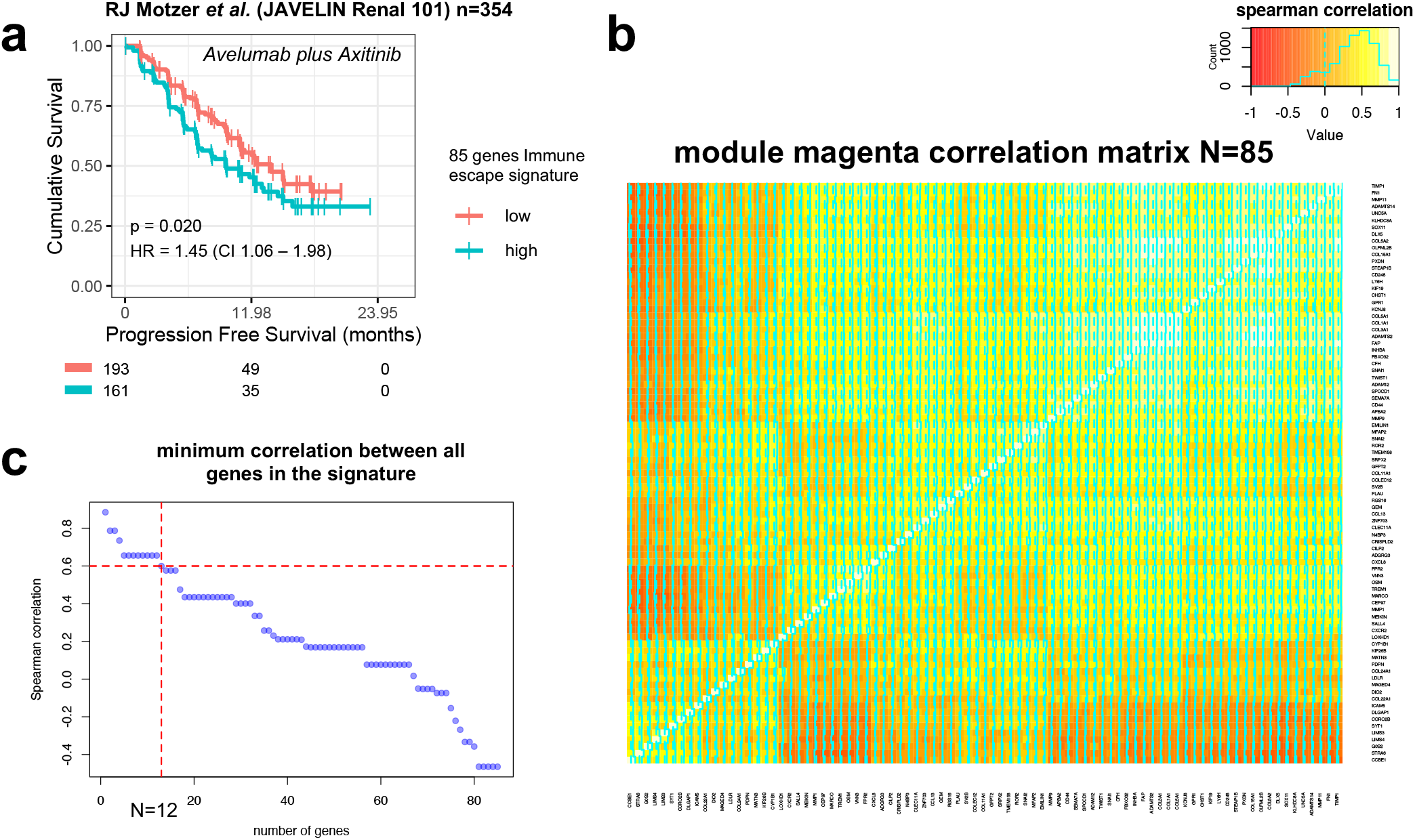
Refinement of immune escape gene signature. **a)** 85 genes immune escape gene signature is strongly associated with response to Avelumab plus Axitinib in JAVELIN Renal 101 trial. **b)** Pairwise spearman correlation between 85 genes in module 16 (immune escape). **c)** Refinement of 85 genes into 12 genes with the highest pairwise spearman correlation.

## Supplementary Materials

### Supplementary Tables

**Table S1:** Data availability.

**Table S2:** Patient characteristics and relevant clinical data.

**Table S3:** WTS related gene signatures.

**Table S4:** ITH classification.

## References

1. Liu X-D, Hoang A, Zhou L, Kalra S, Yetil A, Sun M, Ding Z, Zhang X, Bai S, German P: Resistance to antiangiogenic therapy is associated with an immunosuppressive tumor microenvironment in metastatic renal cell carcinoma. Cancer immunology research 2015, 3:1017–1029.

2. Rooney MS, Shukla SA, Wu CJ, Getz G, Hacohen N: Molecular and genetic properties of tumors associated with local immune cytolytic activity. Cell 2015, 160:48–61.

3. Şenbabaoğlu Y, Gejman RS, Winer AG, Liu M, Van Allen EM, de Velasco G, Miao D, Ostrovnaya I, Drill E, Luna A: Tumor immune microenvironment characterization in clear cell renal cell carcinoma identifies prognostic and immunotherapeutically relevant messenger RNA signatures. Genome biology 2016, 17:1–25.

4. Snyder A, Makarov V, Merghoub T, Yuan J, Zaretsky JM, Desrichard A, Walsh LA, Postow MA, Wong P, Ho TS: Genetic basis for clinical response to CTLA-4 blockade in melanoma. New England Journal of Medicine 2014, 371:2189–2199.

5. Le DT, Durham JN, Smith KN, Wang H, Bartlett BR, Aulakh LK, Lu S, Kemberling H, Wilt C, Luber BS: Mismatch repair deficiency predicts response of solid tumors to PD-1 blockade. Science 2017, 357:409–413.

6. Samstein RM, Lee C-H, Shoushtari AN, Hellmann MD, Shen R, Janjigian YY, Barron DA, Zehir A, Jordan EJ, Omuro A: Tumor mutational load predicts survival after immunotherapy across multiple cancer types. Nature genetics 2019, 51:202–206.

7. Havel JJ, Chowell D, Chan TA: The evolving landscape of biomarkers for checkpoint inhibitor immunotherapy. Nature Reviews Cancer 2019, 19:133–150.

8. McDermott DF, Huseni MA, Atkins MB, Motzer RJ, Rini BI, Escudier B, Fong L, Joseph RW, Pal SK, Reeves JA: Clinical activity and molecular correlates of response to atezolizumab alone or in combination with bevacizumab versus sunitinib in renal cell carcinoma. Nature medicine 2018, 24:749–757.

9. Braun DA, Hou Y, Bakouny Z, Ficial M, Sant’Angelo M, Forman J, Ross-Macdonald P, Berger AC, Jegede OA, Elagina L: Interplay of somatic alterations and immune infiltration modulates response to PD-1 blockade in advanced clear cell renal cell carcinoma. Nature medicine 2020, 26:909–918.

10. Motzer RJ, Robbins PB, Powles T, Albiges L, Haanen JB, Larkin J, Mu XJ, Ching KA, Uemura M, Pal SK: Avelumab plus axitinib versus sunitinib in advanced renal cell carcinoma: Biomarker analysis of the phase 3 JAVELIN Renal 101 trial. Nature medicine 2020, 26:1733–1741.

11. Motzer RJ, Escudier B, McDermott DF, George S, Hammers HJ, Srinivas S, Tykodi SS, Sosman JA, Procopio G, Plimack ER: Nivolumab versus everolimus in advanced renal-cell carcinoma. New England Journal of Medicine 2015, 373:1803–1813.

12. Yarchoan M, Hopkins A, Jaffee EM: Tumor mutational burden and response rate to PD-1 inhibition. The New England journal of medicine 2017, 377:2500.

13. Rini BI, Plimack ER, Stus V, Gafanov R, Hawkins R, Nosov D, Pouliot F, Alekseev B, Soulières D, Melichar B: Pembrolizumab plus axitinib versus sunitinib for advanced renal-cell carcinoma. New England Journal of Medicine 2019, 380:1116–1127.

14. Turajlic S, Xu H, Litchfield K, Rowan A, Horswell S, Chambers T, O’Brien T, Lopez JI, Watkins TB, Nicol D: Deterministic evolutionary trajectories influence primary tumor growth: TRACERx renal. Cell 2018, 173:595–610. e511.

15. Krishna C, DiNatale RG, Kuo F, Srivastava RM, Vuong L, Chowell D, Gupta S, Vanderbilt C, Purohit TA, Liu M: Single-cell sequencing links multiregional immune landscapes and tissue-resident T cells in ccRCC to tumor topology and therapy efficacy. Cancer Cell 2021.

16. Au L, Hatipoglu E, de Massy MR, Litchfield K, Beattie G, Rowan A, Schnidrig D, Thompson R, Byrne F, Horswell S: Determinants of anti-PD-1 response and resistance in clear cell renal cell carcinoma. Cancer cell 2021, 39:1497–1518. e1411.

17. Dobin A, Davis CA, Schlesinger F, Drenkow J, Zaleski C, Jha S, Batut P, Chaisson M, Gingeras TR: STAR: ultrafast universal RNA-seq aligner. Bioinformatics 2013, 29:15–21.

18. Lawrence M, Huber W, Pages H, Aboyoun P, Carlson M, Gentleman R, Morgan MT, Carey VJ: Software for computing and annotating genomic ranges. PLoS Comput Biol 2013, 9:e1003118.

19. Karolchik D, Baertsch R, Diekhans M, Furey TS, Hinrichs A, Lu Y, Roskin KM, Schwartz M, Sugnet CW, Thomas DJ: The UCSC genome browser database. Nucleic acids research 2003, 31:51–54.

20. Love MI, Huber W, Anders S: Moderated estimation of fold change and dispersion for RNA-seq data with DESeq2. Genome biology 2014, 15:1–21.

21. Yoshihara K, Shahmoradgoli M, Martínez E, Vegesna R, Kim H, Torres-Garcia W, Treviño V, Shen H, Laird PW, Levine DA: Inferring tumour purity and stromal and immune cell admixture from expression data. Nature communications 2013, 4:1–11.

22. Barbie DA, Tamayo P, Boehm JS, Kim SY, Moody SE, Dunn IF, Schinzel AC, Sandy P, Meylan E, Scholl C: Systematic RNA interference reveals that oncogenic KRAS-driven cancers require TBK1. Nature 2009, 462:108–112.

23. Newman AM, Liu CL, Green MR, Gentles AJ, Feng W, Xu Y, Hoang CD, Diehn M, Alizadeh AA: Robust enumeration of cell subsets from tissue expression profiles. Nature methods 2015, 12:453–457.

24. Bindea G, Mlecnik B, Tosolini M, Kirilovsky A, Waldner M, Obenauf AC, Angell H, Fredriksen T, Lafontaine L, Berger A: Spatiotemporal dynamics of intratumoral immune cells reveal the immune landscape in human cancer. Immunity 2013, 39:782–795.

25. Golkaram M, Salmans ML, Kaplan S, Vijayaraghavan R, Martins M, Khan N, Garbutt C, Wise A, Yao J, Casimiro S: HERVs establish a distinct molecular subtype in stage II/III colorectal cancer with poor outcome. NPJ genomic medicine 2021, 6:1–11.

26. Li H, Durbin R: Fast and accurate short read alignment with Burrows–Wheeler transform. bioinformatics 2009, 25:1754–1760.

27. McKenna A, Hanna M, Banks E, Sivachenko A, Cibulskis K, Kernytsky A, Garimella K, Altshuler D, Gabriel S, Daly M: The Genome Analysis Toolkit: a MapReduce framework for analyzing next-generation DNA sequencing data. Genome research 2010, 20:1297–1303.

28. DePristo MA, Banks E, Poplin R, Garimella KV, Maguire JR, Hartl C, Philippakis AA, Del Angel G, Rivas MA, Hanna M: A framework for variation discovery and genotyping using next-generation DNA sequencing data. Nature genetics 2011, 43:491.

29. Koboldt DC, Zhang Q, Larson DE, Shen D, McLellan MD, Lin L, Miller CA, Mardis ER, Ding L, Wilson RK: VarScan 2: somatic mutation and copy number alteration discovery in cancer by exome sequencing. Genome research 2012, 22:568–576.

30. Kim S, Scheffler K, Halpern AL, Bekritsky MA, Noh E, Källberg M, Chen X, Kim Y, Beyter D, Krusche P: Strelka2: fast and accurate calling of germline and somatic variants. Nature methods 2018, 15:591–594.

31. Rimmer A, Phan H, Mathieson I, Iqbal Z, Twigg SR, Wilkie AO, McVean G, Lunter G: Integrating mapping-, assembly-and haplotype-based approaches for calling variants in clinical sequencing applications. Nature genetics 2014, 46:912–918.

32. Larson DE, Harris CC, Chen K, Koboldt DC, Abbott TE, Dooling DJ, Ley TJ, Mardis ER, Wilson RK, Ding L: SomaticSniper: identification of somatic point mutations in whole genome sequencing data. Bioinformatics 2012, 28:311–317.

33. Ellrott K, Bailey MH, Saksena G, Covington KR, Kandoth C, Stewart C, Hess J, Ma S, Chiotti KE, McLellan M: Scalable open science approach for mutation calling of tumor exomes using multiple genomic pipelines. Cell systems 2018, 6:271–281. e277.

34. McLaren W, Gil L, Hunt SE, Riat HS, Ritchie GR, Thormann A, Flicek P, Cunningham F: The ensembl variant effect predictor. Genome biology 2016, 17:1–14.

35. Amemiya HM, Kundaje A, Boyle AP: The ENCODE blacklist: identification of problematic regions of the genome. Scientific reports 2019, 9:1–5.

36. Shen R, Seshan VE: FACETS: allele-specific copy number and clonal heterogeneity analysis tool for high-throughput DNA sequencing. Nucleic acids research 2016, 44:e131–e131.

37. McGranahan N, Rosenthal R, Hiley CT, Rowan AJ, Watkins TB, Wilson GA, Birkbak NJ, Veeriah S, Van Loo P, Herrero J: Allele-specific HLA loss and immune escape in lung cancer evolution. Cell 2017, 171:1259–1271. e1211.

38. Paradis E, Claude J, Strimmer K: APE: analyses of phylogenetics and evolution in R language. Bioinformatics 2004, 20:289–290.

39. Raynaud F, Mina M, Tavernari D, Ciriello G: Pan-cancer inference of intra-tumor heterogeneity reveals associations with different forms of genomic instability. PLoS genetics 2018, 14:e1007669.

40. Wu TD, Madireddi S, de Almeida PE, Banchereau R, Chen Y-JJ, Chitre AS, Chiang EY, Iftikhar H, O’Gorman WE, Au-Yeung A: Peripheral T cell expansion predicts tumour infiltration and clinical response. Nature 2020, 579:274–278.

41. Nazarov V: immunarch. bot & Eugene Rumynskiy. immunomind/immunarch: 0.6. 5: Basic single-cell support. Zenodo; 2020.

42. Horn HS: Measurement of” overlap” in comparative ecological studies. The American Naturalist 1966, 100:419–424.

43. Chowell D, Krishna C, Pierini F, Makarov V, Rizvi NA, Kuo F, Morris LG, Riaz N, Lenz TL, Chan TA: Evolutionary divergence of HLA class I genotype impacts efficacy of cancer immunotherapy. Nature medicine 2019, 25:1715–1720.

44. Langfelder P, Horvath S: WGCNA: an R package for weighted correlation network analysis. BMC bioinformatics 2008, 9:1–13.

45. Chakravarthy A, Khan L, Bensler NP, Bose P, De Carvalho DD: TGF-β-associated extracellular matrix genes link cancer-associated fibroblasts to immune evasion and immunotherapy failure. Nature communications 2018, 9:1–10.

46. Turajlic S, Xu H, Litchfield K, Rowan A, Chambers T, Lopez JI, Nicol D, O’Brien T, Larkin J, Horswell S: Tracking cancer evolution reveals constrained routes to metastases: TRACERx renal. Cell 2018, 173:581–594. e512.

47. Hakimi AA, Voss MH, Kuo F, Sanchez A, Liu M, Nixon BG, Vuong L, Ostrovnaya I, Chen Y-B, Reuter V: Transcriptomic profiling of the tumor microenvironment reveals distinct subgroups of clear cell renal cell cancer: data from a randomized phase III trial. Cancer discovery 2019, 9:510–525.

48. Horn S, Leonardelli S, Sucker A, Schadendorf D, Griewank KG, Paschen A: Tumor CDKN2A-associated JAK2 loss and susceptibility to immunotherapy resistance. JNCI: Journal of the National Cancer Institute 2018, 110:677–681.

49. Han G, Yang G, Hao D, Lu Y, Thein K, Simpson BS, Chen J, Sun R, Alhalabi O, Wang R: 9p21 loss confers a cold tumor immune microenvironment and primary resistance to immune checkpoint therapy. Nature communications 2021, 12:1–19.

50. Laydon DJ, Bangham CR, Asquithp B: Estimating T-cell repertoire diversity: limitations of classical estimators and a new approach. Philosophical Transactions of the Royal Society B: Biological Sciences 2015, 370:20140291.

51. Riley TP, Keller GL, Smith AR, Davancaze LM, Arbuiso AG, Devlin JR, Baker BM: Structure based prediction of neoantigen immunogenicity. Frontiers in immunology 2019, 10:2047.

52. Bonaventura P, Alcazer V, Mutez V, Tonon L, Martin J, Chuvin N, Michel E, Boulos RE, Estornes Y, Valladeau-Guilemond J: Identification of shared tumor epitopes from endogenous retroviruses inducing high-avidity cytotoxic T cells for cancer immunotherapy. Science Advances 2022, 8:eabj3671.

53. Panda A, de Cubas AA, Stein M, Riedlinger G, Kra J, Mayer T, Smith CC, Vincent BG, Serody JS, Beckermann KE: Endogenous retrovirus expression is associated with response to immune checkpoint blockade in clear cell renal cell carcinoma. JCI insight 2018, 3.

54. Smith CC, Beckermann KE, Bortone DS, De Cubas AA, Bixby LM, Lee SJ, Panda A, Ganesan S, Bhanot G, Wallen EM: Endogenous retroviral signatures predict immunotherapy response in clear cell renal cell carcinoma. The Journal of clinical investigation 2019, 128:4804–4820.

55. Nargund AM, Pham CG, Dong Y, Wang PI, Osmangeyoglu HU, Xie Y, Aras O, Han S, Oyama T, Takeda S: The SWI/SNF protein PBRM1 restrains VHL-loss-driven clear cell renal cell carcinoma. Cell reports 2017, 18:2893–2906.

56. Hakimi AA, Attalla K, DiNatale RG, Ostrovnaya I, Flynn J, Blum KA, Ged Y, Hoen D, Kendall SM, Reznik E: A pan-cancer analysis of PBAF complex mutations and their association with immunotherapy response. Nature communications 2020, 11:1–11.

57. Liu X-D, Kong W, Peterson CB, McGrail DJ, Hoang A, Zhang X, Lam T, Pilie PG, Zhu H, Beckermann KE: PBRM1 loss defines a nonimmunogenic tumor phenotype associated with checkpoint inhibitor resistance in renal carcinoma. Nature communications 2020, 11:1–14.

58. Zhou M, Leung JY, Gessner KH, Hepperla AJ, Simon JM, Davis IJ, Kim WY: PBRM1 inactivation promotes upregulation of human endogenous retroviruses in a HIF-dependent manner. Cancer immunology research 2022.

59. Zhang Y, Narayanan SP, Mannan R, Raskind G, Wang X, Vats P, Su F, Hosseini N, Cao X, Kumar-Sinha C: Single-cell analyses of renal cell cancers reveal insights into tumor microenvironment, cell of origin, and therapy response. Proceedings of the National Academy of Sciences 2021, 118.

60. Zhang AW, McPherson A, Milne K, Kroeger DR, Hamilton PT, Miranda A, Funnell T, Little N, de Souza CP, Laan S: Interfaces of malignant and immunologic clonal dynamics in ovarian cancer. Cell 2018, 173:1755–1769. e1722.

61. Levine JH, Simonds EF, Bendall SC, Davis KL, El-ad DA, Tadmor MD, Litvin O, Fienberg HG, Jager A, Zunder ER: Data-driven phenotypic dissection of AML reveals progenitor-like cells that correlate with prognosis. Cell 2015, 162:184–197.

62. Xu C, Su Z: Identification of cell types from single-cell transcriptomes using a novel clustering method. Bioinformatics 2015, 31:1974–1980.

63. Hegde PS, Chen DS: Top 10 challenges in cancer immunotherapy. Immunity 2020, 52:17–35.

64. Motzer RJ, Banchereau R, Hamidi H, Powles T, McDermott D, Atkins MB, Escudier B, Liu L-F, Leng N, Abbas AR: Molecular subsets in renal cancer determine outcome to checkpoint and angiogenesis blockade. Cancer Cell 2020, 38:803–817. e804.

65. O’Donnell JS, Teng MW, Smyth MJ: Cancer immunoediting and resistance to T cell-based immunotherapy. Nature reviews Clinical oncology 2019, 16:151–167.

